# Signs and symptoms preceding cancer diagnosis: a data-driven analysis of primary care records

**DOI:** 10.64898/2026.10.20.26366010

**Authors:** Danielle Newby, Elin Rowlands, Edward Burn, Francesc Xavier Avilés-Jurado, Àlvar Roselló Serrano, Leonidas Mavroeidis, Antonella Delmestri, Eng Hooi Tan, Daniel Prieto Alhambra

## Abstract

**Background:** Earlier cancer diagnosis improves outcomes, yet non-specific clinical presentations can make cancer recognition challenging in primary care.

**Aim:** To characterise shared and site-specific patterns of signs, symptoms, and conditions recorded in UK primary care before diagnosis of 23 solid cancers.

**Design and setting:** Population-based matched case control study using Clinical Practice Research Datalink (CPRD) Gold. Adults aged ≥18 years with incident cancer were matched 1:1 by age and sex to individuals without cancer of interest.

**Method:** Signs, symptoms, and conditions recorded 365 to 31 days before diagnosis were compared in cases versus controls using standardised mean differences (SMDs). Clinical codes with SMDs >0.1 were classified as red (>0.4), orange (>0.2 to ≤0.4), or yellow (>0.1 to ≤0.2) flags.

**Results:** Overall, 382,813 cases were matched to site-specific cancer-free controls. Overall, 561 clinical codes had SMDs >0.1. Leading red flags before diagnosis were dysphagia in oesophageal cancer (SMD 0.63), sore throat in hypopharyngeal cancer (0.60), severe dyskaryosis in cervical cancer (0.57), rectal haemorrhage in rectal cancer (0.57), and breast lump in breast cancer (0.52). Abdominal pain was imbalanced across 12 cancers and constipation and dysphagia each across eight, with flag classifications varying by site.

**Conclusion:** Data-driven characterisation identified established red flags and broader pre-diagnostic clinical features. Shared signs, symptoms and conditions varied in flag classification between cancers, revealing cancer-specific signatures and providing a foundation for evaluating combinations of clinical features for earlier cancer recognition in primary care.

**How this fits in:** Established red flags for individual cancers are well recognised but less is known about the broader spectrum of signs, symptoms, and conditions recorded before cancer diagnosis in primary care and how these vary across cancers. This study characterised pre-diagnostic signs, symptoms and conditions across 23 solid cancers, identifying established red flags alongside a broader range of recorded features. Several signs, symptoms, and conditions occurred across multiple cancers but differed in flag classification between cancer sites, revealing distinct cancer-specific clinical signatures. These findings highlight the complexity of cancer presentation and provide a foundation for future research into data-driven approaches to support earlier cancer recognition in primary care.

## INTRODUCTION

Earlier diagnosis and treatment of cancer is essential for improving survival(1), yet many cancers continue to be diagnosed at advanced stages(2) despite modest improvements in recent years(3). Efforts to improve earlier cancer diagnosis are complex and depend on timely patient presentation, appropriate clinical assessment in primary care, and efficient diagnostic pathways within the healthcare system(4).

Despite advances in screening programmes and diagnostic technologies, most patients will continue to be diagnosed after development of symptoms over the coming decade(5,6). Consequently, primary care clinicians will remain central to recognising and responding to potential cancer signs and symptoms to facilitate timely diagnosis(7). However, many cancers present with non-specific clinical features that are easily attributed to benign conditions or other underlying health problems(8–10).

Several studies have identified red-flag symptoms associated with individual cancers, such as rectal bleeding in colorectal cancer(11) and haemoptysis, cough and dyspnoea in lung cancer(12), informing referral guidelines and risk prediction tools(13,14). Recent advances in the availability of large-scale electronic health records (EHR), analytical methods and computational power offer opportunities for data-driven discovery of red flags preceding cancer diagnosis across multiple cancer types. A recent phenome-wide study using EHR from the All of Us Research Program demonstrated the potential of this approach to characterise clinical concepts preceding cancer diagnosis(15). However, results from volunteer research cohorts may not fully reflect routine clinical practice, potentially limiting the generalisability of these findings. Applying a large-scale, data-driven approach to routinely collected UK primary care data provides an opportunity to characterise routinely recorded signs, symptoms and conditions preceding cancer diagnosis and identify both shared and cancer-specific patterns of presentation.

The aim of this study was to perform a large-scale, data-driven characterisation of routinely recorded signs, symptoms and conditions during the year preceding diagnosis across 23 solid cancers in UK primary care to describe cancer-shared and cancer-specific patterns of pre-diagnostic presentation.

## METHODS

### Study design, setting, and data sources

We conducted a population-based matched case-control study using UK primary care data from the Clinical Practice Research Datalink (CPRD) Gold (January 2025)(16). CPRD Gold contains pseudonymized patient-level information on demographics, lifestyle data, clinical diagnoses, prescriptions, and preventive care provided to patients(17) and CPRD Gold was standardised and harmonised to the Observational Medical Outcomes Partnership (OMOP) Common Data Model (CDM)(18).

### Study participants

Eligible participants were aged ≥18 years, had a recorded diagnosis of the cancer of interest, and at least one year of prior observation in CPRD Gold to ensure adequate capture of recorded signs, symptoms and conditions preceding diagnosis. Twenty-three solid cancer sites were included: anus, bladder, brain, breast, cervix, colon, corpus uteri, esophagus, hypopharynx, kidney, larynx, lip and oral cavity, liver, lung, nasopharynx, oropharynx, ovary, pancreas, prostate, rectum, salivary gland, stomach and thyroid.

To reduce inclusion of prevalent cancers, individuals with a record of any other cancer (apart from skin cancer) during the year preceding the diagnosis of the cancer of interest were excluded. For sex-specific cancers, individuals whose recorded sex was inconsistent with the cancer site were also excluded. Each cancer case was matched 1:1 to an individual with at least one year of prior observation without the cancer of interest or with any other cancer diagnosed in the prior year based on year of birth and sex. The index date for matched individuals was assigned as the diagnosis date of the corresponding cancer case. Matched individuals were selected randomly without replacement within each cancer-specific cohort. Individuals could contribute to more than one cancer-specific cohort, either as a case or matched individual, provided they independently met the eligibility criteria for each cohort.

### Outcome definition

Cancer phenotypes were developed using a standardised phenotyping reproducible framework(19). We focused on incident solid tumours excluding benign neoplasms, in situ neoplasms, recurrent cancers, secondary malignancies, skin cancers, and lymphomas. OMOP-based computable phenotypes and analytical code are publicly available on GitHub (https://github.com/oxford-pharmacoepi/CancerSignsSymptoms).

### Statistical methods

Baseline characteristics of cancer patients and matched individuals were summarised using predefined comorbid conditions(20). Continuous variables were summarised using medians and interquartile ranges (IQRs), and categorical variables were reported using counts and percentages.

The primary pre-diagnostic window was defined as 365 to 31 days before the index date to minimise capture of the immediate diagnostic episode while capturing the broader pre-diagnostic period. A wider window of 365 to 91 days was used in sensitivity analysis.

Large-scale characterisation was performed separately for each cancer site by comparing cancer cases with matched individuals without the cancer of interest. All signs, symptoms and conditions recorded in the OMOP CDM Condition domain during the pre-diagnostic window were evaluated using standardised OMOP concepts (hereafter, concepts). Each concept represents a distinct clinical entity, such as a specific diagnosis, sign or symptom, and provides a standardised representation of clinical information recorded using different source codes.

For each concept, the proportion of individuals with at least one recorded occurrence was calculated in each group for cancer patients and matched individuals with absent concepts assigned zero. Differences between groups were quantified using standardised mean difference (SMD). An SMD >0.1 was used as a threshold to identify potentially meaningful differences between groups(21). For ease of interpretation and visualisation, concepts with SMD >0.4, >0.2 to ≤0.4 and >0.1 to ≤0.2 were pragmatically classified as red, orange and yellow flag groups, respectively. These study defined categories were used descriptively to distinguish concepts according to the magnitude of observed differences and do not represent established clinical thresholds. Concepts were additionally mapped to higher-level clinical presentation groups adapted from Zakkak *et al* (5). Group assignment was based on the clinical meaning of each concept, supported by its position within the OMOP concept hierarchy, with each concept assigned to the most clinically relevant presentation group for visualisation.

Results based on fewer than five cases were suppressed to minimise the risk of patient identification. Analyses were performed using R version 4.2.3.

## RESULTS

A total of 382,813 individuals with at least one of 23 cancer sites were matched 1-to-1 to individuals without the specific cancer of interest (total population = 765,626) (**Table 1**). The largest cohorts consisted of breast (n = 92,456), prostate (n = 69,543) and lung cancer (n = 56,415) patients, and the smallest ones were nasopharyngeal (n = 414) and hypopharyngeal cancer (n = 690) patients. Median age ranged from 49 years for cervical cancer to 75 years for stomach cancer. Detailed baseline characteristics are presented in **Supplementary Table S1.**

**Table 1.** Baseline characteristics of patients by cancers site and age and sex matched individuals.

| Cancer | Cohort<sup>a</sup> | N | Age<br>[Q25 - Q75] | Female<br>N% |
| --- | --- | --- | --- | --- |
| Anus | Cancer | 1,319 | 67 [57 - 76] | 855 (64.82%) |
|  | Matched |  |  |  |
| Bladder | Cancer | 22,124 | 74 [66 - 81] | 6,117 (27.65%) |
|  | Matched |  |  |  |
| Brain | Cancer | 7,936 | 64 [53 - 74] | 3,493 (44.01%) |
|  | Matched |  |  |  |
| Breast | Cancer | 92,456 | 62 [52 - 73] | 91,924 (99.42%) |
|  | Matched |  |  |  |
| Cervix | Cancer | 4,574 | 49 [37 - 64] | 4,574 (100.00%) |
|  | Matched |  |  |  |
| Colon | Cancer | 34,485 | 73 [64 - 80] | 16,010 (46.43%) |
|  | Matched |  |  |  |
| Corpus uteri | Cancer | 8,623 | 67 [59 - 75] | 8,623 (100.00%) |
|  | Matched |  |  |  |
| Esophagus | Cancer | 15,634 | 72 [63 - 79] | 4,999 (31.98%) |
|  | Matched |  |  |  |
| Hypopharynx | Cancer | 690 | 67 [60 - 76] | 163 (23.62%) |
|  | Matched |  |  |  |
| Kidney | Cancer | 5,843 | 68 [59 - 77] | 2,219 (37.98%) |
|  | Matched |  |  |  |
| Larynx | Cancer | 2,899 | 67 [59 - 74] | 574 (19.80%) |
|  | Matched |  |  |  |
| Lip/oral cavity | Cancer | 3,634 | 64 [56 - 74] | 1,408 (38.75%) |
|  | Matched |  |  |  |
| Liver | Cancer | 5,069 | 71 [63 - 78] | 1,480 (29.20%) |
|  | Matched |  |  |  |
| Lung | Cancer | 56,415 | 72 [65 - 79] | 26,304 (46.63%) |
|  | Matched |  |  |  |
| Nasopharynx | Cancer | 414 | 61 [52 - 72] | 136 (32.85%) |
|  | Matched |  |  |  |
| Oropharynx | Cancer | 2,613 | 61 [54 - 69] | 668 (25.56%) |
|  | Matched |  |  |  |
| Ovary | Cancer | 9,064 | 66 [56 - 75] | 9,064 (100.00%) |
|  | Matched |  |  |  |
| Pancreas | Cancer | 11,114 | 73 [64 - 80] | 5,576 (50.17%) |
|  | Matched |  |  |  |
| Prostate | Cancer | 69,543 | 72 [65 - 78] | - |
|  | Matched |  |  |  |
| Rectum | Cancer | 16,960 | 71 [62 - 79] | 6,580 (38.80%) |
|  | Matched |  |  |  |
| Salivary gland | Cancer | 814 | 68 [55 - 79] | 335 (41.15%) |
|  | Matched |  |  |  |
| Stomach | Cancer | 7,673 | 75 [67 - 82] | 2,912 (37.95%) |
|  | Matched |  |  |  |
| Thyroid | Cancer | 2,917 | 52 [39 - 67] | 2,138 (73.29%) |
|  | Matched |  |  |  |

<sup>a</sup> Cancer cases were matched to individuals without the cancer of interest by year of birth and sex

Across 23 cancer sites, 561 concepts had SMD > 0.1, ranging from 64 for nasopharyngeal cancer to two for breast cancer. The largest SMD were for dysphagia in esophageal cancer (SMD 0.63), sore throat in hypopharyngeal cancer (0.60), rectal haemorrhage in rectal cancer (0.57), and severe dyskaryosis in cervical cancer (0.57).

Several features occurred across multiple cancers but differed in flag classification. Abdominal pain occurred across 12 cancers, with red flags for pancreatic and ovarian cancers and orange flag classification for colon, liver, stomach and kidney cancers.

Constipation and dysphagia each occurred across eight cancers while cough, iron deficiency anaemia, sore throat, swelling, lymphadenopathy and neck mass each occurred across seven. Blood pressure findings occurred across 20 cancers but were predominantly classified within the orange and yellow flag. All concepts with SMD >0.1, and their flag classifications are provided in **Supplementary Table S2**.

Grouping concepts into presentation domains showed that cancers within the same anatomical systems generally shared presentation patterns (**Figure 1**). Upper abdominal, lower abdominal and non-specific domains occurred across multiple cancers, while respiratory presentations predominated across head and neck and lung cancers, while genitourinary and female-specific cancers showed predominantly organ-specific presentation. Despite similarities within anatomical groups, distinct cancer-specific patterns were evident (**Figures 2-4** and **Supplementary Figures S1-S4**).

**Figure 1.**
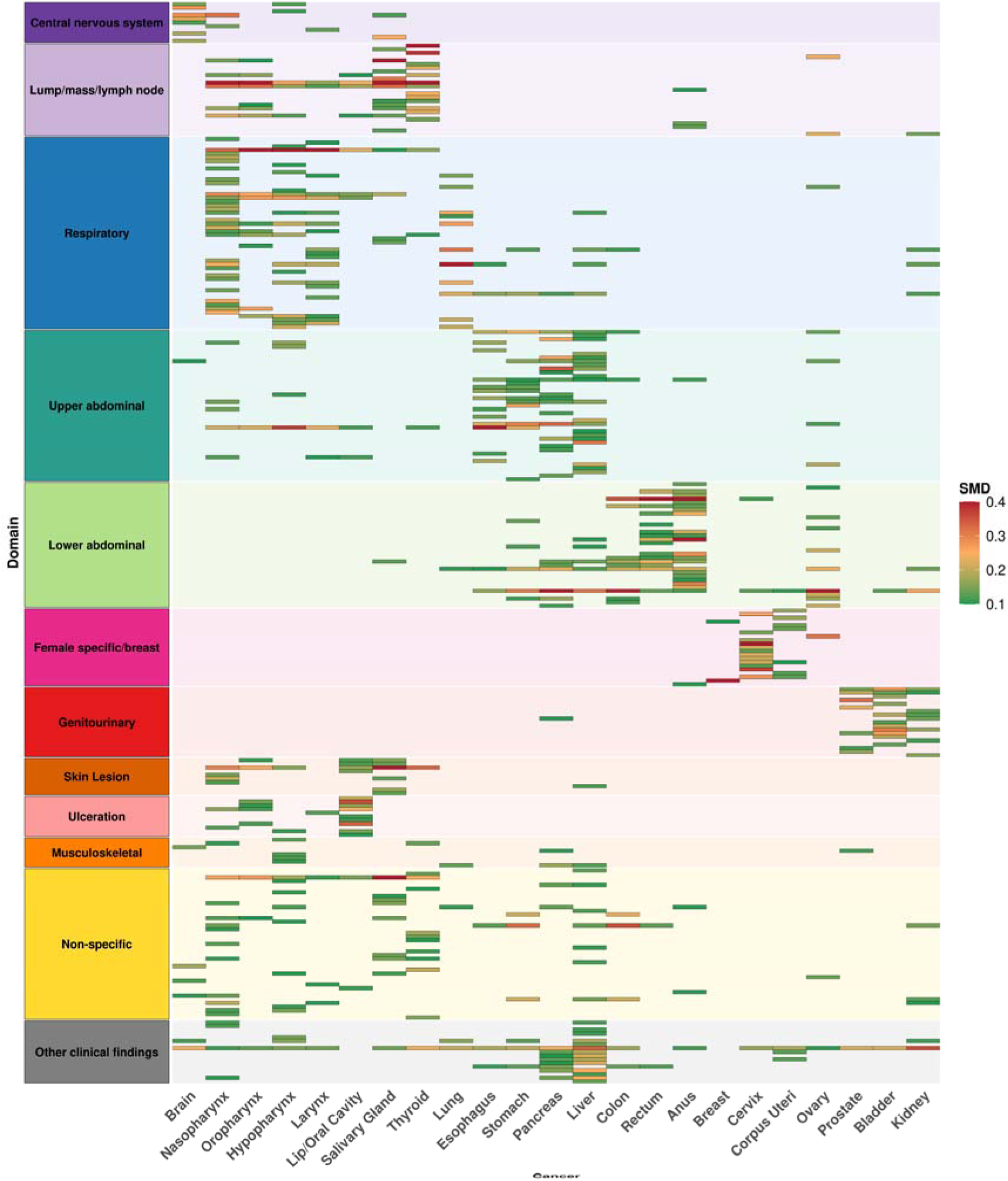
Heatmap showing signs, symptoms and conditions with absolute standardized mean differences (SMD) greater than 0.1 across specific cancer sites, grouped by presentation domain. Colours represent the magnitude of the SMD, with higher values indicating greater differences between cases and matched individuals.

**Figure 2.**
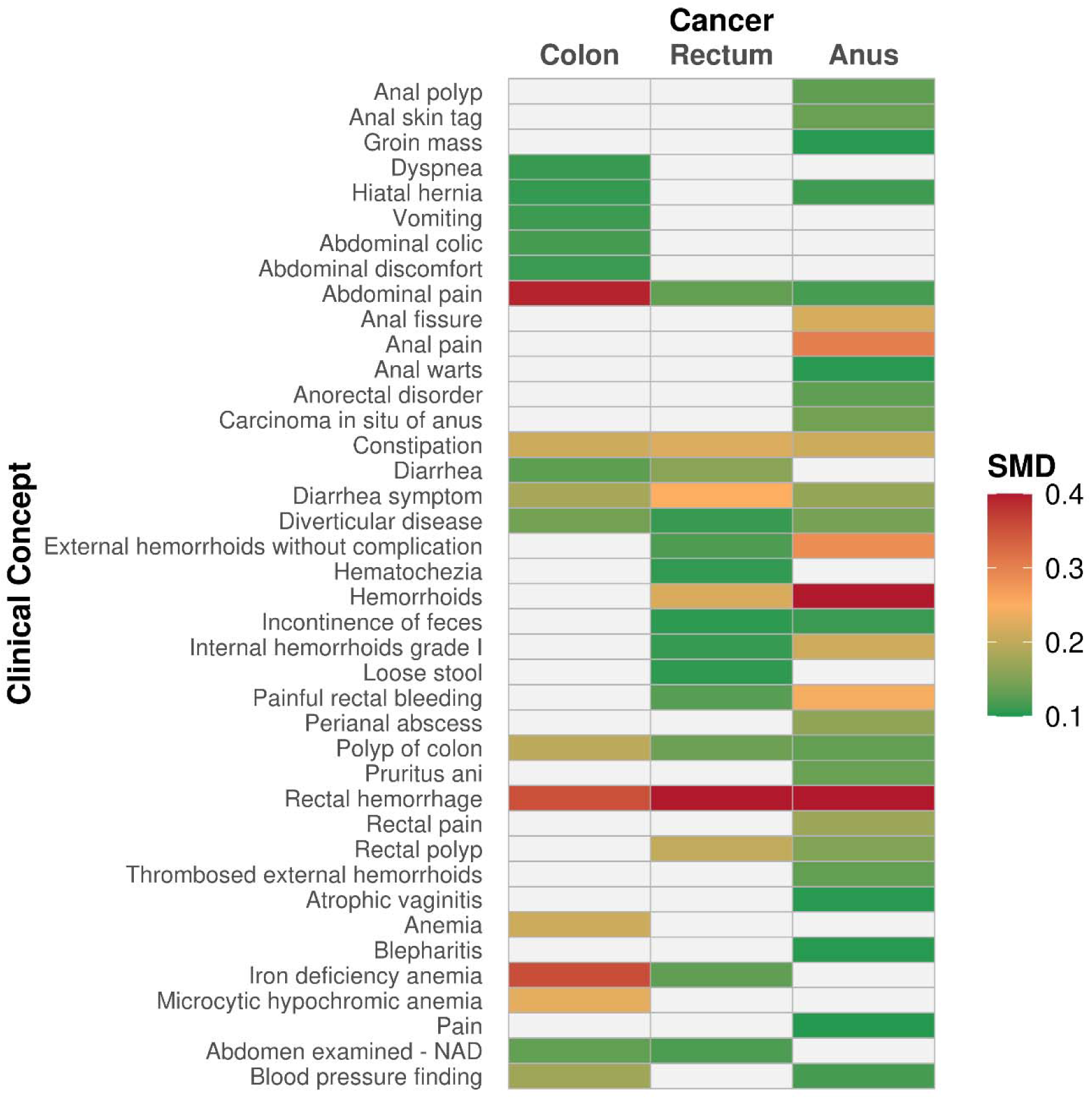
Heatmap showing site-specific signs, symptoms and condition concepts with absolute standardized mean differences (SMD) greater than 0.1 for lower gastrointestinal cancers. Colours represent the magnitude of the SMD, with higher values indicating greater differences between cases and matched individuals. NAD; No Abnormality Detected.

Lower gastrointestinal cancers shared predominantly lower abdominal and anorectal features (**Figure 2**). Rectal haemorrhage was a red flag for rectal (SMD 0.57) and anal cancers (0.54), but an orange flag for colon cancer (0.35). Abdominal pain (0.39) and iron deficiency anaemia (0.36) were orange flags for colon cancer, while anal cancer showed anorectal-specific features including haemorrhoids as a red flag (0.51) and anal pain (0.30) as an orange flag. Diarrhoea, polyp of colon and diverticular disease were predominantly yellow flags across these cancers.

Upper gastrointestinal cancers shared several abdominal concepts (**Figure 3**). Abdominal pain ranged from a yellow flag group for esophageal cancer (SMD 0.16) to the red flag for pancreatic cancer (0.53), with orange flags for stomach (0.31) and liver cancer (0.32). Dysphagia was the only red flag for esophageal cancer (0.63) and an orange flag for stomach cancer (0.23). Other prominent concepts included iron deficiency anaemia for stomach cancer, jaundice for pancreatic cancer and cirrhosis for liver cancer. Liver cancer had more concepts with SMDs > 0.1 than other cancers shown in **Figure 3**.

**Figure 3.**
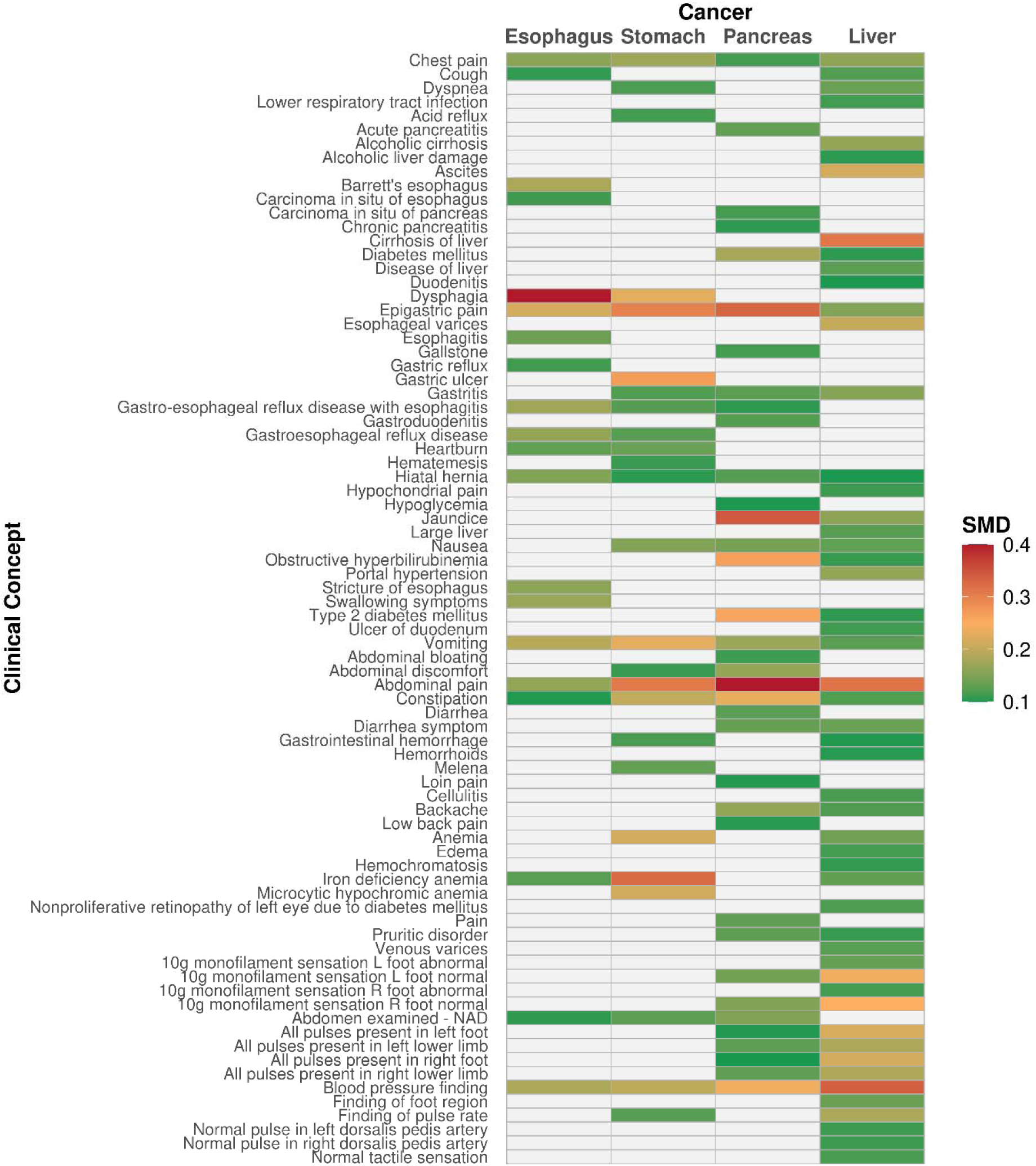
Heatmap showing site-specific signs, symptoms and condition concepts with absolute standardized mean differences (SMD) greater than 0.1 for upper gastrointestinal cancers. Colours represent the magnitude of the SMD, with higher values indicating greater differences between cases and matched individuals. NAD; No Abnormality Detected.

Head and neck cancers overlapped with respiratory and head and neck presentations (**Figure 4**). Sore throat was a red flag for hypopharyngeal (SMD 0.60), oropharyngeal (0.53) and laryngeal cancers (0.47), but an orange flag for nasopharyngeal cancer (0.33). Mass of neck was a red flag for oropharyngeal (0.51), salivary gland (0.43) and nasopharyngeal cancers (0.43), and an orange flag for hypopharyngeal cancer (0.27). Lip/oral cavity cancers had orange flags, including mouth ulcers (0.36) and disorder of tongue (0.34).

**Figure 4.**
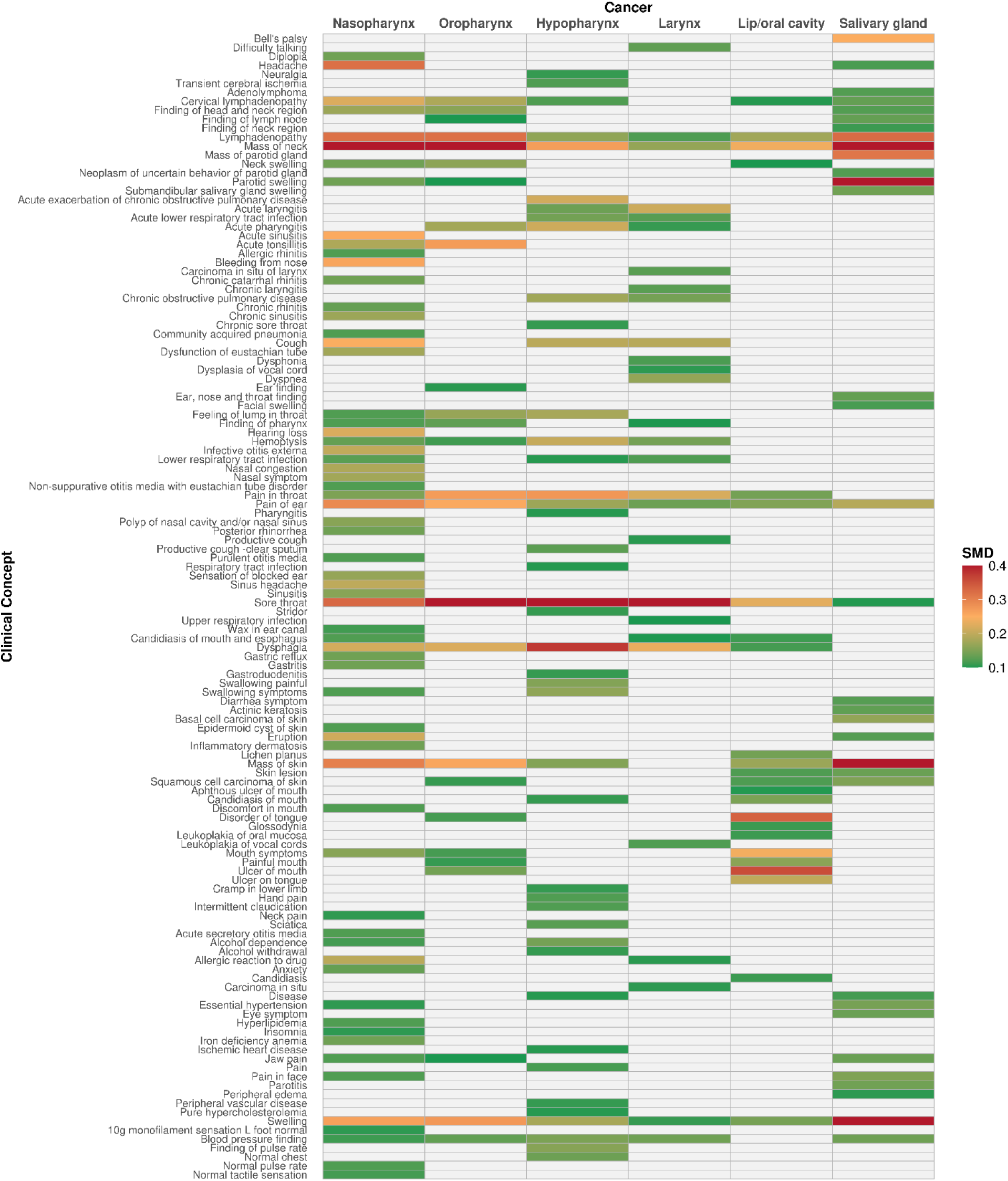
Heatmap showing site-specific signs, symptoms and condition concepts with absolute standardized mean differences (SMD) greater than 0.1 for head and neck cancers. Colours represent the magnitude of the SMD, with higher values indicating greater differences between cases and matched individuals.

Other cancers demonstrated predominantly organ-specific patterns (**Supplementary Figures S1-S4**). Lung cancer was characterised by respiratory concepts, with cough as the only red flag (SMD 0.40). Genitourinary concepts dominated in prostate and bladder cancer, including prostatism (SMD 0.33) and nocturia (0.23) for prostate cancer and frank haematuria (0.31) and urinary tract infectious disease (0.29) for bladder cancer.

Kidney cancer had no red flags, with acquired renal cystic disease (0.15), loin pain (0.14), constipation (0.14), and cough (0.13) among identified concepts. Breast lump was the only red flag for breast cancer (SMD 0.52), and severe dyskaryosis (0.57) for cervical cancer. Abdominal pain was the only red flag for ovarian cancer (0.51) with cyst of ovary (0.32), pelvic mass (0.24), constipation (0.24), and abdominal distension (0.21) as orange flags. Corpus uteri cancer had no red or orange flags, with polyp of the corpus uteri (0.17) and uterine leiomyoma (0.16) as the highest-ranking yellow flags.

Brain cancer had no red flags, with headache (SMD 0.29) and seizure (0.28) ranking the highest. Thyroid cancer had red flags for neck mass (0.54) and thyroid nodules (0.46).

In sensitivity analysis, excluding the 90 days before diagnosis (365-to-91 days) reduced the number of concepts with SMD > 0.1 from 561 to 299, and the number of red flags from 20 to 1 and orange flags from 123 to 56. Mass of neck remained a red flag for thyroid cancer (SMD 0.41), while nasopharyngeal cancer had the greatest number of orange flags remaining. Despite the attenuation using this window, patterns across anatomical cancer groups remained broadly consistent with the primary analysis (**Supplementary Table S3**).

## DISCUSSION

### Summary

In this large UK primary care study, we identified a wide range of signs, symptoms and conditions recorded during the year preceding incident cancer diagnosis that differed between individuals with cancer and matched individuals without the cancer of interest. Many red flags reflected recognised cancer presentations, while orange and yellow flags in addition captured a broader spectrum of cancer shared and cancer-specific pre-diagnostic features. Notably, several features occurred across multiple cancers but differed in flag classification between cancer sites, resulting in distinct cancer-specific patterns. Differences were attenuated after excluding the 90 days before diagnosis, although patterns of presentation persisted.

### Strengths and limitations

Strengths of this study include the use of a large UK primary care database to systematically evaluate recorded signs, symptoms and conditions preceding diagnosis across multiple cancer sites. Analysing individual cancers separately, enabled the identification of cancer-shared and cancer-specific patterns of presentation, while the use of age- and sex-matched individuals allowed differentiation of concepts occurring disproportionately before cancer diagnosis. Unlike retrospective audit studies relying on clinician-reported symptoms(5), routinely recorded EHR can reduce recall bias, avoiding retrospective assignment of presenting symptoms.

Several limitations should be considered. Individuals were not matched on healthcare utilization because increasing healthcare contact may form part of the pathway to cancer diagnosis; matching on this could have resulted in overmatching.

Although excluding the 30 days preceding diagnosis reduced capture of the immediate diagnostic episode, some recorded concepts may reflect investigations and/or increased healthcare contact rather than early manifestations of cancer. The choice of exclusion window is therefore important when distinguishing early features from events related to the diagnostic process. We addressed this in a sensitivity analysis excluding the 90 days preceding diagnosis to assess the robustness of our findings to a longer exclusion period. Furthermore, cancers may be diagnosed in secondary care before being communicated to general practice(22) therefore recorded diagnosis date in CPRD Gold can occur after the true diagnosis date and some clinical concepts potentially might reflect the diagnostic process or early management. Clinical concepts were evaluated individually rather than in broader phenotypes containing multiple concepts; variation in coding practices may have dispersed similar presentations across related concepts, reducing observed differences for individual concepts. Finally, the study restricted to the OMOP Condition domain; consequently, clinically relevant presenting features recorded in the Observation domain, including fatigue, weight findings, hoarseness and gynecological bleeding were not captured by our analysis.

### Comparison with existing literature

Abdominal pain, dysphagia and constipation were among the most frequently observed concepts across multiple cancer types, although the SMD magnitude varied considerably between cancer sites(23,24). As these are also common presentations of non-malignant disease in primary care, (25) our findings highlight the challenge clinicians face, in distinguishing the small proportion of patients with common, non-specific symptoms who subsequently receive a cancer diagnosis(26). This diagnostic uncertainty is a recognised contributor to delayed cancer diagnoses and poorer outcomes.(27)

Many red flags corresponded to established cancer presentations, including dysphagia before diagnosis of esophageal cancer(28), breast lump before breast cancer(15,29), hematuria before bladder cancer(30), jaundice before pancreatic cancer(31,32) and rectal hemorrhage before colorectal cancers(11). These findings are consistent with observational studies(5,33,34), referral guidance (NICE NG12)(14), risk prediction tools(13,35) and other data-driven analyses(15), supporting the clinical plausibility of our data-driven approach. Identified red flags also included diagnosed conditions potentially reflecting comorbidity, diagnostic overlap, or alternative diagnoses considered before cancer diagnosis. Future studies using clinically relevant comparator groups, rather than the general population, could help distinguish features associated with cancer from those conditions with overlapping presentations.

Our study additionally demonstrated that the breadth and pattern of recorded concepts differed between cancer sites. Some cancers showed focused site-specific presentations such as breast and bladder cancer, whereas ovarian and upper gastrointestinal cancers were characterised by more heterogeneous clinical presentations(5,33,36,37). Head and neck cancers also demonstrated both recognised red flags and a wider range of respiratory- and neck-related presentations(38,39).

These findings suggest that, for many cancer sites, the pre-diagnostic period is characterised by multiple recorded clinical features rather than a single dominant presentation(40,41) Not all identified concepts represented direct manifestations of cancer. Blood pressure and other clinical findings across multiple cancers may partly reflect increasing assessment and healthcare contact during the diagnostic pathway, consistent with previous evidence of rising consultation frequency before cancer diagnosis (42). This suggests that the pre-diagnostic period reflects not only manifestations of cancer but also increasing clinical interactions. Supporting this interpretation, excluding the 90 days preceding diagnosis reduced both the number and magnitude of differences, indicating that these were concentrated closer to diagnosis. Nevertheless, broader patterns persisted, indicating that differences in recorded clinical information extend beyond the period immediately preceding diagnosis.

### Implications for research and/or practice

Established red flags remain central for cancer recognition and referral in primary care(13,14), but our findings demonstrate that the pre-diagnostic period encompasses a broader spectrum of recorded clinical features, with patterns differing between cancer sites. For clinicians, these findings highlight the complexity and heterogeneity of cancer presentation in primary care, particularly where common, non-specific features occur across multiple cancers(43,44). Future studies should evaluate whether data-driven approaches using routinely collected healthcare data have the potential to support and provide additional value alongside existing referral pathways and risk prediction tools for earlier cancer recognition(45).

In conclusion, this large data-driven characterisation of recorded signs, symptoms and conditions identified established cancer red flags alongside broader cancer-shared and cancer-specific patterns of recorded clinical features preceding diagnosis. Together, these findings provide a description of the recorded clinical information preceding cancer diagnosis in UK primary care and illustrate the value of data-driven analyses for understanding the complexity of the pre-diagnostic period and provide a foundation for future research to improve earlier cancer recognition.

## ADDITIONAL INFORMATION

### Author Contributions

Conceptualization; DN, DPA; Data harmonisation and data quality assessment; AD; Formal analysis; DN; Funding acquisition; DPA; Supervision; DPA; Interpretation of results: All authors; Roles/Writing - original draft: DN; Writing - review & editing: All authors.

### Data availability statement

This study is based in part on data from the Clinical Practice Research Datalink (CPRD) Gold obtained under licence from the UK Medicines and Healthcare products Regulatory Agency. The data is provided by patients and collected by the NHS as part of their care and support. The interpretation and conclusions contained in this study are those of the author/s alone. Patient level data used in this study were obtained through an approved CPRD protocol (application number 24_004724 https://www.cprd.com/approved-studies/time-trends-prevalence-incidence-survival-and-characterisation-cancers-united) and are only available following an approval process to safeguard the confidentiality of patient data. Details on how to apply for data access can be found at https://www.cprd.com/access-data. Individual patient consent was not required, as CPRD data are de-identified and provided under ethical approval from the UK Health Research Authority (HRA) and the NHS Health and Social Care Research Ethics Committee.

## Acknowledgements

The authors thank Hezekiah Omulo and Mandickel Kamtengeni for database standardization and database technical support. DN would also like to acknowledge her late husband, Sam Francis, who took a particular interest in the findings of this research and with whom she had many useful discussions. She dedicates this paper to his memory.

## REFERENCES

1. Hawkes N. Cancer survival data emphasise importance of early diagnosis. BMJ. 2019 Jan 25;364:l408. doi:10.1136/BMJ.L408 PubMed PMID: 30683652.

2. Crosby D, Bhatia S, Brindle KM, Coussens LM, Dive C, Emberton M, et al. Early detection of cancer. Science (1979). 2022 Mar 18;375(6586). doi:10.1126/SCIENCE.AAY9040;ISSUE:ISSUE:DOI PubMed PMID: 35298272.

3. NHS England:» Early cancer diagnosis in England reaches highest ever level [Internet]. [cited 2026 Jul 7]. Available from: https://www.england.nhs.uk/2025/01/early-cancer-diagnosis-in-england-reaches-highest-ever-level/

4. Raleigh V, Goldblatt P, Colombo F. Ethnic differences in mortality in England and Wales after the pandemic. BMJ. 2025. doi:10.1136/BMJ.Q2856 PubMed PMID: 39746709.

5. Zakkak N, Barclay ME, Swann R, McPhail S, Rubin G, Abel GA, et al. The presenting symptom signatures of incident cancer: evidence from the English 2018 National Cancer Diagnosis Audit. British Journal of Cancer 2023 130:2. 2023 Dec 6;130(2):297–307. doi:10.1038/s41416-023-02507-4 PubMed PMID: 38057397.

6. Richards MA, Hiom S, Hamilton W. Diagnosing cancer earlier: what progress is being made? Br J Cancer. 2023 Feb 2;128(3):441. doi:10.1038/S41416-023-02171-8 PubMed PMID: 36725918.

7. Rubin G, Berendsen A, Crawford SM, Dommett R, Earle C, Emery J, et al. The expanding role of primary care in cancer control. Lancet Oncol. 2015 Sep 1;16(12):1231–72. doi:10.1016/S1470-2045(15)00205-3 PubMed PMID: 26431866.

8. Forster AS, Renzi C, Lyratzopoulos G. Diagnosing cancer in patients with ‘non-alarm’ symptoms: Learning from diagnostic care innovations in Denmark. Cancer Epidemiol. 2018 Jun 1;54:101–3. doi:10.1016/j.canep.2018.03.011 PubMed PMID: 29702426.

9. Lyratzopoulos G, Vedsted P, Singh H. Understanding missed opportunities for more timely diagnosis of cancer in symptomatic patients after presentation. Br J Cancer. 2015;112 Suppl 1(Suppl 1):S84–91. doi:10.1038/BJC.2015.47 PubMed PMID: 25734393.

10. Langue G, Pinker I, Moran V, Pilleron S. Attributing non-specific symptoms to cancer in general practice: A scoping review. PLoS One. 2025 Jun 1;20(6):e0322264. doi:10.1371/JOURNAL.PONE.0322264 PubMed PMID: 40549694.

11. Bekkink OM, McCowan C, Falk GA, Teljeur C, Van De Laar FA, Fahey T. Diagnostic accuracy systematic review of rectal bleeding in combination with other symptoms, signs and tests in relation to colorectal cancer. British Journal of Cancer 2010 102:1. 2009 Nov 24;102(1):48–58. doi:10.1038/sj.bjc.6605426 PubMed PMID: 19935790.

12. Prado MG, Kessler LG, Au MA, Burkhardt HA, Zigman Suchsland M, Kowalski L, et al. Symptoms and signs of lung cancer prior to diagnosis: case–control study using electronic health records from ambulatory care within a large US-based tertiary care centre. BMJ Open. 2023 Apr 1;13(4):e068832. doi:10.1136/BMJOPEN-2022-068832 PubMed PMID: 37080616.

13. Hippisley-Cox J, Coupland C. Development and validation of risk prediction algorithms to estimate future risk of common cancers in men and women: prospective cohort study. BMJ Open. 2015;5(3):e007825. doi:10.1136/BMJOPEN-2015-007825 PubMed PMID: 25783428.

14. Overview | Suspected cancer: recognition and referral | Guidance | NICE.

15. Christian C, Rich D, Bang EJ, Bair AB, Richardson BE, Millington JL, et al. Phenome-Wide Association Study of Pre-Cancer Diagnosis Electronic Health Records Identifies Risk and Inverse Associations in the All of Us Research Program. medRxiv. 2026 May 28;2026.05.26.26353823. doi:10.64898/2026.05.26.26353823

16. Page: CPRD GOLD January 2025 | CPRD [Internet]. [cited 2026 Jul 7]. Available from: https://www.cprd.com/doi/cprd-gold-january-2025

17. Sanchez-Santos MT, Axson EL, Dedman D, Delmestri A. Data Resource Profile Update: CPRD GOLD. Int J Epidemiol. 2025 Jun 11;54(4). doi:10.1093/IJE/DYAF077 PubMed PMID: 40499193.

18. Voss EA, Makadia R, Matcho A, Ma Q, Knoll C, Schuemie M, et al. Feasibility and utility of applications of the common data model to multiple, disparate observational health databases. J Am Med Inform Assoc. 2015 May 1;22(3):553–64. doi:10.1093/JAMIA/OCU023 PubMed PMID: 25670757.

19. Dernie F, Corby G, Robinson A, Bezer J, Mercade-Besora N, Griffier R, et al. Standardised and Reproducible Phenotyping Using Distributed Analytics and Tools in the Data Analysis and Real World Interrogation Network (DARWIN EU). Pharmacoepidemiol Drug Saf. 2024 Nov 1;33(11). doi:10.1002/PDS.70042 PubMed PMID: 39532529.

20. López-Sánchez I, Palomar-Cros A, Claire R, Pérez-Crespo L, Giuliodori A, Koblbauer I, et al. Comorbidities, medication use, and overall survival in eight cancers: a multinational cohort study of 1.7 million patients across Europe. The Lancet Regional Health – Europe. 2026 Apr 1;63:101585. doi:10.1016/J.LANEPE.2025.101585 PubMed PMID: 41631168.

21. Austin PC. Balance diagnostics for comparing the distribution of baseline covariates between treatment groups in propensity-score matched samples. Stat Med. 2009 Nov 10;28(25):3083–107. doi:10.1002/SIM.3697 PubMed PMID: 19757444.

22. Whitfield E, White B, Barclay ME, Rafiq M, Renzi C, Rous B, et al. Differences in recording of cancer diagnosis between datasets in England: A population-based study of linked cancer registration, hospital, and primary care data. Cancer Epidemiol. 2025 Feb 1;94:102703. doi:10.1016/J.CANEP.2024.102703 PubMed PMID: 39612750.

23. Koo MM, von Wagner C, Abel GA, McPhail S, Hamilton W, Rubin GP, et al. The nature and frequency of abdominal symptoms in cancer patients and their associations with time to help-seeking: evidence from a national audit of cancer diagnosis. J Public Health (Oxf). 2018 Sep 1;40(3):e388. doi:10.1093/PUBMED/FDX188 PubMed PMID: 29385513.

24. Wilkins T, Gillies RA, Thomas AM, Wagner PJ. The prevalence of dysphagia in primary care patients: a HamesNet Research Network study. J Am Board Fam Med. 2007 Mar;20(2):144–50. doi:10.3122/JABFM.2007.02.060045 PubMed PMID: 17341750.

25. Finley CR, Chan DS, Garrison S, Korownyk C, Kolber MR, Campbell S, et al. What are the most common conditions in primary care? Systematic review. Canadian Family Physician. 2018 Nov 1;64(11):832. PubMed PMID: 30429181.

26. Hamilton W. Cancer diagnosis in primary care. British Journal of General Practice. 2010 Feb 1;60(571):121–8. doi:10.3399/BJGP10X483175 PubMed PMID: 20132704.

27. Neal RD, Tharmanathan P, France B, Din NU, Cotton S, Fallon-Ferguson J, et al. Is increased time to diagnosis and treatment in symptomatic cancer associated with poorer outcomes? Systematic review. Br J Cancer. 2015;112 Suppl 1(Suppl 1):S92–107. doi:10.1038/BJC.2015.48 PubMed PMID: 25734382.

28. Humphrys E, Walter FM, Rubin G, Emery JD, Johnson M, Richards A, et al. Patient symptom experience prior to a diagnosis of oesophageal or gastric cancer: a multi-methods study. BJGP Open. 2020;4(1):bjgpopen20X101001. doi:10.3399/BJGPOPEN20X101001 PubMed PMID: 31911419.

29. Koo MM, von Wagner C, Abel GA, McPhail S, Rubin GP, Lyratzopoulos G. Typical and atypical presenting symptoms of breast cancer and their associations with diagnostic intervals: Evidence from a national audit of cancer diagnosis. Cancer Epidemiol. 2017 Jun 1;48:140–6. doi:10.1016/J.CANEP.2017.04.010 PubMed PMID: 28549339.

30. Shephard EA, Stapley S, Neal RD, Rose P, Walter FM, Hamilton WT. Clinical features of bladder cancer in primary care. The British Journal of General Practice. 2012;62(602):e598. doi:10.3399/BJGP12X654560 PubMed PMID: 22947580.

31. Liao W, Clift AK, Patone M, Coupland C, Izquierdo AG, Pereira SP, et al. Identifying symptoms associated with diagnosis of pancreatic exocrine and neuroendocrine neoplasms: a nested case-control study of the UK primary care population. British Journal of General Practice. 2021 Nov 1;71(712):e836–45. doi:10.3399/BJGP.2021.0153 PubMed PMID: 34544691.

32. Ceniceros L, Prados S, Gallego RA. Jaundice in Pancreatic Cancer. Textbook of Pancreatic Cancer: Principles and Practice of Surgical Oncology. 2021 Jan 1;1215–25. doi:10.1007/978-3-030-53786-9_78/TABLES/3

33. Bosch X, Montori-Palacin E, Cadena-Rutllan M, Mota-Gomes T, Montoya M, Moreno P, et al. Symptomatic presentation of cancer in patients diagnosed following emergency self-referral and prior general practice consultations. Postgrad Med. 2026 May 19. doi:10.1080/00325481.2026.2674374

34. Hamilton W. The CAPER studies: five case-control studies aimed at identifying and quantifying the risk of cancer in symptomatic primary care patients. Br J Cancer. 2009 Dec 3;101(Suppl 2):S80. doi:10.1038/SJ.BJC.6605396 PubMed PMID: 19956169.

35. Hippisley-Cox J, Coupland CA. Development and external validation of prediction algorithms to improve early diagnosis of cancer. Nature Communications 2025 16:1. 2025 May 7;16(1):3660-. doi:10.1038/s41467-025-57990-5 PubMed PMID: 40335498.

36. Koo MM, Hamilton W, Walter FM, Rubin GP, Lyratzopoulos G. Symptom Signatures and Diagnostic Timeliness in Cancer Patients: A Review of Current Evidence. Neoplasia (United States). 2018 Feb 1;20(2):165–74. doi:10.1016/j.neo.2017.11.005 PubMed PMID: 29253839.

37. Bankhead CR, Kehoe ST, Austoker J. Symptoms associated with diagnosis of ovarian cancer: a systematic review. BJOG. 2005 Jul;112(7):857–65. doi:10.1111/J.1471-0528.2005.00572.X PubMed PMID: 15957984.

38. Bradley PT, Lee YK, Albutt A, Hardman J, Kellar I, Odo C, et al. Nomenclature of the symptoms of head and neck cancer: a systematic scoping review. Front Oncol. 2024;14:1404860. doi:10.3389/FONC.2024.1404860 PubMed PMID: 38952557.

39. Minhas S, Sajjad A, Kashif M, Taj F, Waddani H Al, Khurshid Z. Oral Ulcers Presentation in Systemic Diseases: An Update. Open Access Maced J Med Sci. 2019 Oct 15;7(19):3341. doi:10.3889/OAMJMS.2019.689 PubMed PMID: 31949540.

40. Koo MM, Swann R, McPhail S, Abel GA, Elliss-Brookes L, Rubin GP, et al. Presenting symptoms of cancer and stage at diagnosis: evidence from a cross-sectional, population-based study. Lancet Oncol. 2020 Jan 1;21(1):73–9. doi:10.1016/S1470-2045(19)30595-9 PubMed PMID: 31704137.

41. Avilés-Jurado FX, Valduvieco I, Medrano-Martorell S, Muxí A, Lop J, Cuesta F, et al. Comprehensive cancer centre accreditation: experience and achievements of a head neck cancer unit. Acta Otorrinolaringologica (English Edition). 2026 Jun 12;512371. doi:10.1016/J.OTOENG.2026.512371

42. White B, Renzi C, Rafiq M, Abel GA, Jensen H, Lyratzopoulos G. Does changing healthcare use signal opportunities for earlier detection of cancer? A review of studies using information from electronic patient records. Cancer Epidemiol. 2022 Feb 1;76:102072. doi:10.1016/J.CANEP.2021.102072 PubMed PMID: 34876377.

43. Rubin G, Berendsen A, Crawford SM, Dommett R, Earle C, Emery J, et al. The expanding role of primary care in cancer control. Lancet Oncol. 2015 Sep 1;16(12):1231–72. doi:10.1016/S1470-2045(15)00205-3 PubMed PMID: 26431866.

44. Moore SF, Abel G, Mounce LTA. Challenges of diagnosing rare cancers in primary care. The British Journal of General Practice. 2024 Aug 1;74(745):340. doi:10.3399/BJGP24X738789 PubMed PMID: 39054086.

45. National Cancer Plan for England:: delivering world class cancer care [Internet]. 2026 [cited 2026 Jul 7]. Available from: https://www.gov.uk/government/publications/national-cancer-plan-for-england

