## Supplement 2 for "Signs and symptoms preceding cancer diagnosis: a data-driven analysis of primary care records"

[Table S2: Clinical concepts with absolute standardised mean differences (SMDs) > 0.1 for window 365-31 days before index showing percentages of each concept in cancer patients and matched individuals with the designation into flag group (RED SMD > 0.4, ORANGE SMD > 0.2 and <= 0.4, YELLOW SMD >0.1 and <= 0.2) and classification each presentation domain adapted from Zakkak et al (2024)	1](#_Toc409360094)

[Table S3: Clinical concepts with absolute standardised mean differences (SMDs) > 0.1 for window 365-91 days before index showing percentages of each concept in cancer patients and matched individuals with the designation into flag group (RED SMD > 0.4, ORANGE SMD > 0.2 and <= 0.4, YELLOW SMD >0.1 and <= 0.2) and classification each presentation domain adapted from Zakkak et al (2024)	28](#_Toc1942951286)

### **Table S2: Clinical concepts with absolute standardised mean differences (SMDs) > 0.1 for window 365-31 days before index showing percentages of each concept in cancer patients and matched individuals with the designation into flag group (RED SMD > 0.4, ORANGE SMD > 0.2 and <= 0.4, YELLOW SMD >0.1 and <= 0.2) and classification each presentation domain adapted from Zakkak *et al* (2024)**

| **Cancer** | **Code Name (Code ID)** | **Cancer %** | **Matched %** | **SMD** | **Assigned Domain** | **Assigned Flag** |
| --- | --- | --- | --- | --- | --- | --- |
| Anus | Abdominal pain (200219) | 3.71 | 1.82 | 0.1155 | Lower abdominal | YELLOW |
|  | Anal fissure (76725) | 2.73 | 0.15 | 0.2178 | Lower abdominal | ORANGE |
|  | Anal pain (442764) | 4.4 | 0 | 0.3034 | Lower abdominal | ORANGE |
|  | Anal polyp (4228686) | 0.83 | 0 | 0.1294 | Lump/mass/lymph node | YELLOW |
|  | Anal skin tag (4112182) | 0.91 | 0 | 0.1355 | Lump/mass/lymph node | YELLOW |
|  | Anal warts (4084948) | 0.53 | 0 | 0.1032 | Lower abdominal | YELLOW |
|  | Anorectal disorder (4145825) | 0.83 | 0 | 0.1294 | Lower abdominal | YELLOW |
|  | Atrophic vaginitis (201078) | 0.53 | 0 | 0.1032 | Female specific/breast | YELLOW |
|  | Blepharitis (378425) | 0.53 | 0 | 0.1032 | Non-specific | YELLOW |
|  | Blood pressure finding (4214962) | 58.76 | 53.07 | 0.1148 | Other clinical findings | YELLOW |
|  | Carcinoma in situ of anus (78110) | 0.99 | 0 | 0.1414 | Lower abdominal | YELLOW |
|  | Constipation (75860) | 6.22 | 2.05 | 0.2106 | Lower abdominal | ORANGE |
|  | Diarrhea symptom (4145808) | 3.26 | 0.91 | 0.165 | Lower abdominal | YELLOW |
|  | Diverticular disease (4266809) | 2.2 | 0.53 | 0.1443 | Lower abdominal | YELLOW |
|  | External hemorrhoids without complication (199839) | 4.17 | 0.08 | 0.2865 | Lower abdominal | ORANGE |
|  | Groin mass (4083353) | 0.53 | 0 | 0.1032 | Lump/mass/lymph node | YELLOW |
|  | Hemorrhoids (195562) | 12.51 | 0.45 | 0.5053 | Lower abdominal | RED |
|  | Hiatal hernia (4224004) | 0.83 | 0.08 | 0.1116 | Upper abdominal | YELLOW |
|  | Incontinence of feces (197675) | 1.14 | 0.23 | 0.1105 | Lower abdominal | YELLOW |
|  | Internal hemorrhoids grade I (36715916) | 2.2 | 0 | 0.2121 | Lower abdominal | ORANGE |
|  | Pain (4329041) | 2.73 | 1.29 | 0.1027 | Non-specific | YELLOW |
|  | Painful rectal bleeding (4189790) | 2.88 | 0 | 0.2435 | Lower abdominal | ORANGE |
|  | Perianal abscess (199861) | 1.29 | 0 | 0.1617 | Lower abdominal | YELLOW |
|  | Polyp of colon (4285898) | 1.06 | 0.08 | 0.1305 | Lower abdominal | YELLOW |
|  | Pruritus ani (200169) | 1.29 | 0.15 | 0.1351 | Lower abdominal | YELLOW |
|  | Rectal hemorrhage (4026112) | 13.72 | 0.3 | 0.5448 | Lower abdominal | RED |
|  | Rectal pain (76458) | 1.44 | 0 | 0.1709 | Lower abdominal | YELLOW |
|  | Rectal polyp (4216644) | 1.36 | 0.08 | 0.1518 | Lower abdominal | YELLOW |
|  | Thrombosed external hemorrhoids (201045) | 1.06 | 0.08 | 0.1305 | Lower abdominal | YELLOW |
| Bladder | Abdominal pain (200219) | 3.74 | 1.71 | 0.1249 | Lower abdominal | YELLOW |
|  | Blood pressure finding (4214962) | 71.6 | 61.45 | 0.2163 | Other clinical findings | ORANGE |
|  | Carcinoma in situ of urinary bladder (192855) | 0.83 | 0.01 | 0.1271 | Genitourinary | YELLOW |
|  | Cystitis (195588) | 3.5 | 0.8 | 0.187 | Genitourinary | YELLOW |
|  | Dysuria (197684) | 5.21 | 0.67 | 0.2712 | Genitourinary | ORANGE |
|  | Frank hematuria (75004) | 4.72 | 0.06 | 0.3087 | Genitourinary | ORANGE |
|  | Hematuria syndrome (79864) | 2.03 | 0.05 | 0.1961 | Genitourinary | YELLOW |
|  | Hydronephrosis (433811) | 0.75 | 0.02 | 0.1181 | Genitourinary | YELLOW |
|  | Microscopic hematuria (78472) | 1.84 | 0.04 | 0.1874 | Genitourinary | YELLOW |
|  | Painless hematuria (4059740) | 1.21 | 0 | 0.1565 | Genitourinary | YELLOW |
|  | Recurrent urinary tract infection (4056621) | 1.88 | 0.21 | 0.1648 | Genitourinary | YELLOW |
|  | Retention of urine (192450) | 1.41 | 0.27 | 0.1252 | Genitourinary | YELLOW |
|  | Urinary tract infectious disease (81902) | 7.25 | 1.46 | 0.2866 | Genitourinary | ORANGE |
| Brain | Amnesia (439147) | 2.36 | 0.37 | 0.1721 | Central nervous system | YELLOW |
|  | Anxiety (441542) | 1.71 | 0.59 | 0.1052 | Non-specific | YELLOW |
|  | Blood pressure finding (4214962) | 63.17 | 51.12 | 0.2453 | Other clinical findings | ORANGE |
|  | Cerebrovascular accident (381316) | 2.19 | 0.2 | 0.1839 | Central nervous system | YELLOW |
|  | Collapse (443240) | 1.1 | 0.15 | 0.1208 | Non-specific | YELLOW |
|  | Dizziness (4223938) | 4.04 | 1.25 | 0.1745 | Non-specific | YELLOW |
|  | Dysphasia (441594) | 0.82 | 0.05 | 0.1172 | Central nervous system | YELLOW |
|  | Epilepsy (380378) | 2.92 | 0.11 | 0.2316 | Central nervous system | ORANGE |
|  | Finding of pulse rate (4103199) | 11.53 | 8.17 | 0.1129 | Other clinical findings | YELLOW |
|  | Headache (378253) | 6.84 | 1.26 | 0.2859 | Central nervous system | ORANGE |
|  | Muscle weakness (79908) | 1.3 | 0.11 | 0.1426 | Musculoskeletal | YELLOW |
|  | Nausea (31967) | 1.21 | 0.33 | 0.1008 | Upper abdominal | YELLOW |
|  | Seizure (377091) | 3.91 | 0.08 | 0.2765 | Central nervous system | ORANGE |
|  | Transient cerebral ischemia (373503) | 1.81 | 0.33 | 0.1442 | Central nervous system | YELLOW |
| Breast | Breast lump (80767) | 13.37 | 0.6 | 0.5175 | Female specific/breast | RED |
|  | Pain of breast (73819) | 1.52 | 0.51 | 0.1009 | Female specific/breast | YELLOW |
| Cervix | Abdominal pain (200219) | 5.53 | 3.04 | 0.1232 | Lower abdominal | YELLOW |
|  | Blood pressure finding (4214962) | 54.57 | 46.22 | 0.1676 | Other clinical findings | YELLOW |
|  | Cannot exclude glandular neoplasia on cervical smear (4056545) | 3.83 | 0 | 0.2822 | Female specific/breast | ORANGE |
|  | Carcinoma in situ of uterine cervix (194611) | 6.36 | 0 | 0.3686 | Female specific/breast | ORANGE |
|  | Cervical intraepithelial neoplasia grade 2 (196165) | 0.87 | 0.04 | 0.1236 | Female specific/breast | YELLOW |
|  | Cervical smear - borderline changes (4056546) | 3.26 | 0.46 | 0.2084 | Female specific/breast | ORANGE |
|  | Cervical smear - high grade dyskaryosis (severe) (44810424) | 2.03 | 0 | 0.2036 | Female specific/breast | ORANGE |
|  | Cervical smear - human papillomavirus positive (44810559) | 2.49 | 0.04 | 0.2206 | Female specific/breast | ORANGE |
|  | Cervical smear - mild dyskaryosis (4155377) | 1.33 | 0.22 | 0.1268 | Female specific/breast | YELLOW |
|  | Cervical smear - moderate dyskaryosis (4152694) | 2.38 | 0.04 | 0.2153 | Female specific/breast | ORANGE |
|  | Cervical smear - severe dyskaryosis (4151556) | 14.06 | 0.02 | 0.5707 | Female specific/breast | RED |
|  | Cervical smear result (441138) | 3.8 | 1.18 | 0.1687 | Female specific/breast | YELLOW |
|  | Dyskaryosis on cervical smear (4095039) | 1.05 | 0.02 | 0.1416 | Female specific/breast | YELLOW |
|  | Rectal hemorrhage (4026112) | 1.27 | 0.28 | 0.1131 | Lower abdominal | YELLOW |
|  | Severe dyskaryosis on cervical smear cannot exclude invasive carcinoma (4059358) | 3.41 | 0 | 0.2657 | Female specific/breast | ORANGE |
| Colon | Abdomen examined - NAD (4060865) | 1.75 | 0.41 | 0.1299 | Other clinical findings | YELLOW |
|  | Abdominal colic (4322528) | 0.77 | 0.04 | 0.1151 | Lower abdominal | YELLOW |
|  | Abdominal discomfort (4183041) | 1.14 | 0.23 | 0.1105 | Lower abdominal | YELLOW |
|  | Abdominal pain (200219) | 11.61 | 1.91 | 0.3938 | Lower abdominal | ORANGE |
|  | Anemia (439777) | 3.19 | 0.42 | 0.2092 | Non-specific | ORANGE |
|  | Blood pressure finding (4214962) | 68.13 | 59.82 | 0.1738 | Other clinical findings | YELLOW |
|  | Constipation (75860) | 5.78 | 1.79 | 0.2102 | Lower abdominal | ORANGE |
|  | Diarrhea (196523) | 1.71 | 0.4 | 0.1285 | Lower abdominal | YELLOW |
|  | Diarrhea symptom (4145808) | 4.02 | 1.17 | 0.18 | Lower abdominal | YELLOW |
|  | Diverticular disease (4266809) | 2.52 | 0.74 | 0.1409 | Lower abdominal | YELLOW |
|  | Dyspnea (312437) | 4.69 | 2.64 | 0.1093 | Respiratory | YELLOW |
|  | Hiatal hernia (4224004) | 1.58 | 0.49 | 0.1079 | Upper abdominal | YELLOW |
|  | Iron deficiency anemia (436659) | 7.4 | 0.55 | 0.3561 | Non-specific | ORANGE |
|  | Microcytic hypochromic anemia (4262948) | 2.97 | 0.14 | 0.2302 | Non-specific | ORANGE |
|  | Polyp of colon (4285898) | 2.27 | 0.14 | 0.1962 | Lower abdominal | YELLOW |
|  | Rectal hemorrhage (4026112) | 7.23 | 0.5 | 0.3546 | Lower abdominal | ORANGE |
|  | Vomiting (441408) | 1.66 | 0.51 | 0.1112 | Upper abdominal | YELLOW |
| Corpus Uteri | Abdominal pain (200219) | 4.45 | 2.5 | 0.1066 | Lower abdominal | YELLOW |
|  | All pulses present in left lower limb (37312545) | 4.27 | 2.08 | 0.1251 | Other clinical findings | YELLOW |
|  | All pulses present in right lower limb (37312544) | 4.26 | 2.08 | 0.1247 | Other clinical findings | YELLOW |
|  | Blood pressure finding (4214962) | 65.96 | 57.08 | 0.1833 | Other clinical findings | YELLOW |
|  | Cannot exclude glandular neoplasia on cervical smear (4056545) | 0.79 | 0 | 0.1262 | Female specific/breast | YELLOW |
|  | Carcinoma in situ of endometrium (4241777) | 0.77 | 0.01 | 0.1222 | Female specific/breast | YELLOW |
|  | Cervical smear - borderline changes (4056546) | 0.74 | 0.09 | 0.1012 | Female specific/breast | YELLOW |
|  | Endometrial cystic hyperplasia (200459) | 0.9 | 0 | 0.1348 | Female specific/breast | YELLOW |
|  | Not detected by smear of cervix (4155376) | 8.64 | 5.13 | 0.139 | Female specific/breast | YELLOW |
|  | Polyp of corpus uteri (200779) | 1.51 | 0.05 | 0.1665 | Female specific/breast | YELLOW |
|  | Uterine leiomyoma (197236) | 1.7 | 0.15 | 0.1624 | Female specific/breast | YELLOW |
| Esophagus | Abdomen examined - NAD (4060865) | 1.3 | 0.34 | 0.1066 | Other clinical findings | YELLOW |
|  | Abdominal pain (200219) | 4.54 | 1.7 | 0.1639 | Lower abdominal | YELLOW |
|  | Barrett's esophagus (443344) | 2.03 | 0.14 | 0.1832 | Upper abdominal | YELLOW |
|  | Blood pressure finding (4214962) | 68.06 | 59.3 | 0.1829 | Other clinical findings | YELLOW |
|  | Carcinoma in situ of esophagus (28109) | 0.63 | 0 | 0.1126 | Upper abdominal | YELLOW |
|  | Chest pain (77670) | 4.82 | 1.97 | 0.1579 | Respiratory | YELLOW |
|  | Constipation (75860) | 3.28 | 1.7 | 0.1015 | Lower abdominal | YELLOW |
|  | Cough (254761) | 9.44 | 6.56 | 0.1063 | Respiratory | YELLOW |
|  | Dysphagia (31317) | 17.4 | 0.35 | 0.6284 | Upper abdominal | RED |
|  | Epigastric pain (197381) | 3.36 | 0.42 | 0.2172 | Upper abdominal | ORANGE |
|  | Esophagitis (30753) | 1.29 | 0.13 | 0.1385 | Upper abdominal | YELLOW |
|  | Gastric reflux (4023183) | 1 | 0.15 | 0.1126 | Upper abdominal | YELLOW |
|  | Gastro-esophageal reflux disease with esophagitis (30437) | 2.3 | 0.33 | 0.1736 | Upper abdominal | YELLOW |
|  | Gastroesophageal reflux disease (318800) | 1.68 | 0.13 | 0.1642 | Upper abdominal | YELLOW |
|  | Heartburn (23325) | 1.33 | 0.2 | 0.13 | Upper abdominal | YELLOW |
|  | Hiatal hernia (4224004) | 2.09 | 0.42 | 0.1504 | Upper abdominal | YELLOW |
|  | Iron deficiency anemia (436659) | 1.85 | 0.5 | 0.1255 | Non-specific | YELLOW |
|  | Stricture of esophagus (4268622) | 1.39 | 0.04 | 0.1607 | Upper abdominal | YELLOW |
|  | Swallowing symptoms (4149859) | 1.52 | 0.04 | 0.1688 | Upper abdominal | YELLOW |
|  | Vomiting (441408) | 2.87 | 0.43 | 0.1924 | Upper abdominal | YELLOW |
| Hypopharynx | Acute exacerbation of chronic obstructive pulmonary disease (257004) | 2.61 | 0.14 | 0.2133 | Respiratory | ORANGE |
|  | Acute laryngitis (30133) | 1.3 | 0.14 | 0.1375 | Respiratory | YELLOW |
|  | Acute lower respiratory tract infection (4307774) | 2.75 | 0.87 | 0.1414 | Respiratory | YELLOW |
|  | Acute pharyngitis (25297) | 2.17 | 0 | 0.2106 | Respiratory | ORANGE |
|  | Alcohol dependence (435243) | 1.01 | 0 | 0.1428 | Non-specific | YELLOW |
|  | Alcohol withdrawal (375519) | 0.58 | 0 | 0.108 | Non-specific | YELLOW |
|  | Blood pressure finding (4214962) | 63.19 | 55.94 | 0.1481 | Other clinical findings | YELLOW |
|  | Candidiasis of mouth (29735) | 0.58 | 0 | 0.108 | Ulceration | YELLOW |
|  | Cervical lymphadenopathy (4133459) | 0.72 | 0 | 0.1204 | Lump/mass/lymph node | YELLOW |
|  | Chronic obstructive pulmonary disease (255573) | 1.88 | 0.14 | 0.1747 | Respiratory | YELLOW |
|  | Chronic sore throat (4168068) | 0.58 | 0 | 0.108 | Respiratory | YELLOW |
|  | Cough (254761) | 12.75 | 6.96 | 0.1952 | Respiratory | YELLOW |
|  | Cramp in lower limb (42710045) | 0.58 | 0 | 0.108 | Musculoskeletal | YELLOW |
|  | Disease (4274025) | 0.87 | 0.14 | 0.1031 | Non-specific | YELLOW |
|  | Dysphagia (31317) | 8.12 | 0.58 | 0.3761 | Upper abdominal | ORANGE |
|  | Feeling of lump in throat (4145818) | 1.59 | 0 | 0.1798 | Respiratory | YELLOW |
|  | Finding of pulse rate (4103199) | 14.64 | 9.57 | 0.1559 | Other clinical findings | YELLOW |
|  | Gastroduodenitis (195306) | 0.58 | 0 | 0.108 | Upper abdominal | YELLOW |
|  | Hand pain (4204199) | 0.72 | 0 | 0.1204 | Musculoskeletal | YELLOW |
|  | Hemoptysis (261687) | 2.03 | 0 | 0.2036 | Respiratory | ORANGE |
|  | Intermittent claudication (442774) | 0.72 | 0 | 0.1204 | Musculoskeletal | YELLOW |
|  | Ischemic heart disease (4185932) | 0.87 | 0.14 | 0.1031 | Non-specific | YELLOW |
|  | Lower respiratory tract infection (4175297) | 5.07 | 3.04 | 0.1031 | Respiratory | YELLOW |
|  | Lymphadenopathy (315085) | 1.3 | 0 | 0.1623 | Lump/mass/lymph node | YELLOW |
|  | Mass of neck (4100606) | 3.48 | 0 | 0.2685 | Lump/mass/lymph node | ORANGE |
|  | Mass of skin (4183953) | 1.16 | 0 | 0.1532 | Skin Lesion | YELLOW |
|  | Neuralgia (373852) | 0.58 | 0 | 0.108 | Central nervous system | YELLOW |
|  | Normal chest (3654258) | 1.3 | 0.14 | 0.1375 | Other clinical findings | YELLOW |
|  | Pain (4329041) | 2.32 | 0.87 | 0.1159 | Non-specific | YELLOW |
|  | Pain in throat (259153) | 4.78 | 0.43 | 0.2757 | Respiratory | ORANGE |
|  | Pain of ear (4103476) | 3.04 | 0.72 | 0.1714 | Respiratory | YELLOW |
|  | Peripheral vascular disease (321052) | 0.58 | 0 | 0.108 | Non-specific | YELLOW |
|  | Pharyngitis (4226263) | 0.87 | 0.14 | 0.1031 | Respiratory | YELLOW |
|  | Productive cough -clear sputum (4060224) | 1.16 | 0.14 | 0.1272 | Respiratory | YELLOW |
|  | Pure hypercholesterolemia (437827) | 0.87 | 0.14 | 0.1031 | Non-specific | YELLOW |
|  | Respiratory tract infection (4170143) | 1.16 | 0.29 | 0.1027 | Respiratory | YELLOW |
|  | Sciatica (372409) | 1.45 | 0.29 | 0.1252 | Musculoskeletal | YELLOW |
|  | Sore throat (4147326) | 17.83 | 1.01 | 0.6013 | Respiratory | RED |
|  | Stridor (253321) | 0.58 | 0 | 0.108 | Respiratory | YELLOW |
|  | Swallowing painful (4147218) | 1.16 | 0 | 0.1532 | Upper abdominal | YELLOW |
|  | Swallowing symptoms (4149859) | 1.3 | 0 | 0.1623 | Upper abdominal | YELLOW |
|  | Swelling (4275722) | 2.32 | 0.29 | 0.1796 | Non-specific | YELLOW |
|  | Transient cerebral ischemia (373503) | 0.72 | 0 | 0.1204 | Central nervous system | YELLOW |
| Kidney | Abdominal mass (192438) | 0.96 | 0 | 0.1392 | Lump/mass/lymph node | YELLOW |
|  | Abdominal pain (200219) | 7.36 | 1.93 | 0.2602 | Lower abdominal | ORANGE |
|  | Acquired renal cystic disease (194686) | 1.23 | 0.03 | 0.1521 | Genitourinary | YELLOW |
|  | Allergic reaction to drug (4166257) | 5.19 | 3.13 | 0.1033 | Non-specific | YELLOW |
|  | Anemia (439777) | 1.45 | 0.29 | 0.1252 | Non-specific | YELLOW |
|  | Blood pressure finding (4214962) | 71.74 | 54.85 | 0.3559 | Other clinical findings | ORANGE |
|  | Chest pain (77670) | 3.73 | 1.78 | 0.1193 | Respiratory | YELLOW |
|  | Chronic kidney disease stage 3 (443597) | 2.81 | 1.23 | 0.1125 | Genitourinary | YELLOW |
|  | Constipation (75860) | 3.66 | 1.52 | 0.135 | Lower abdominal | YELLOW |
|  | Cough (254761) | 9.69 | 6.18 | 0.1301 | Respiratory | YELLOW |
|  | Dyspnea (312437) | 4 | 1.99 | 0.1181 | Respiratory | YELLOW |
|  | Finding of pulse rate (4103199) | 14.75 | 11.02 | 0.1115 | Other clinical findings | YELLOW |
|  | Frank hematuria (75004) | 1.54 | 0.09 | 0.1618 | Genitourinary | YELLOW |
|  | Iron deficiency anemia (436659) | 2.11 | 0.5 | 0.1422 | Non-specific | YELLOW |
|  | Loin pain (4152353) | 1.66 | 0.31 | 0.137 | Genitourinary | YELLOW |
|  | Microscopic hematuria (78472) | 0.91 | 0.07 | 0.1205 | Genitourinary | YELLOW |
|  | Neoplasm of uncertain behavior of kidney (198402) | 0.86 | 0 | 0.1317 | Genitourinary | YELLOW |
|  | Retention of urine (192450) | 0.65 | 0.03 | 0.1067 | Genitourinary | YELLOW |
|  | Urinary tract infectious disease (81902) | 3.65 | 1.27 | 0.1541 | Genitourinary | YELLOW |
| Larynx | Acute laryngitis (30133) | 2.24 | 0.03 | 0.2098 | Respiratory | ORANGE |
|  | Acute lower respiratory tract infection (4307774) | 2.66 | 1 | 0.1241 | Respiratory | YELLOW |
|  | Acute pharyngitis (25297) | 1.21 | 0.28 | 0.1083 | Respiratory | YELLOW |
|  | Allergic reaction to drug (4166257) | 4.24 | 2.38 | 0.1041 | Non-specific | YELLOW |
|  | Blood pressure finding (4214962) | 62.09 | 55.23 | 0.1396 | Other clinical findings | YELLOW |
|  | Candidiasis of mouth and esophagus (4142836) | 0.59 | 0.03 | 0.1009 | Upper abdominal | YELLOW |
|  | Carcinoma in situ (433435) | 0.55 | 0 | 0.1052 | Non-specific | YELLOW |
|  | Carcinoma in situ of larynx (24897) | 0.79 | 0 | 0.1262 | Respiratory | YELLOW |
|  | Chronic laryngitis (24970) | 0.79 | 0 | 0.1262 | Respiratory | YELLOW |
|  | Chronic obstructive pulmonary disease (255573) | 2.52 | 0.72 | 0.1429 | Respiratory | YELLOW |
|  | Cough (254761) | 11.8 | 6.31 | 0.1922 | Respiratory | YELLOW |
|  | Difficulty talking (4114720) | 0.83 | 0 | 0.1294 | Central nervous system | YELLOW |
|  | Dysphagia (31317) | 3.1 | 0.17 | 0.2326 | Upper abdominal | ORANGE |
|  | Dysphonia (40326053) | 0.69 | 0 | 0.1179 | Respiratory | YELLOW |
|  | Dysplasia of vocal cord (4147413) | 0.55 | 0 | 0.1052 | Respiratory | YELLOW |
|  | Dyspnea (312437) | 4.59 | 1.72 | 0.1647 | Respiratory | YELLOW |
|  | Finding of pharynx (4022571) | 0.69 | 0.07 | 0.1009 | Respiratory | YELLOW |
|  | Hemoptysis (261687) | 1.28 | 0.1 | 0.1429 | Respiratory | YELLOW |
|  | Leukoplakia of vocal cords (4323991) | 0.72 | 0 | 0.1204 | Ulceration | YELLOW |
|  | Lower respiratory tract infection (4175297) | 5.04 | 2.73 | 0.1198 | Respiratory | YELLOW |
|  | Lymphadenopathy (315085) | 0.97 | 0.1 | 0.1195 | Lump/mass/lymph node | YELLOW |
|  | Mass of neck (4100606) | 1.55 | 0.07 | 0.1657 | Lump/mass/lymph node | YELLOW |
|  | Pain in throat (259153) | 2.97 | 0.24 | 0.2185 | Respiratory | ORANGE |
|  | Pain of ear (4103476) | 2.79 | 0.93 | 0.138 | Respiratory | YELLOW |
|  | Productive cough (4102774) | 1.69 | 0.59 | 0.1038 | Respiratory | YELLOW |
|  | Sore throat (4147326) | 11.42 | 0.62 | 0.4662 | Respiratory | RED |
|  | Swelling (4275722) | 1.21 | 0.28 | 0.1083 | Non-specific | YELLOW |
|  | Upper respiratory infection (4181583) | 3.66 | 2 | 0.1002 | Respiratory | YELLOW |
| Lip/Oral cavity | Aphthous ulcer of mouth (138463) | 0.52 | 0 | 0.1022 | Ulceration | YELLOW |
|  | Candidiasis (433968) | 1.18 | 0.25 | 0.1105 | Non-specific | YELLOW |
|  | Candidiasis of mouth (29735) | 1.24 | 0.06 | 0.1472 | Ulceration | YELLOW |
|  | Candidiasis of mouth and esophagus (4142836) | 0.83 | 0.08 | 0.1116 | Upper abdominal | YELLOW |
|  | Cervical lymphadenopathy (4133459) | 0.61 | 0.03 | 0.1028 | Lump/mass/lymph node | YELLOW |
|  | Disorder of tongue (432941) | 5.72 | 0.14 | 0.3355 | Ulceration | ORANGE |
|  | Dysphagia (31317) | 1.29 | 0.28 | 0.1146 | Upper abdominal | YELLOW |
|  | Glossodynia (441334) | 0.61 | 0 | 0.1108 | Ulceration | YELLOW |
|  | Leukoplakia of oral mucosa (138466) | 0.61 | 0 | 0.1108 | Ulceration | YELLOW |
|  | Lichen planus (132703) | 1.21 | 0.08 | 0.1415 | Skin Lesion | YELLOW |
|  | Lymphadenopathy (315085) | 1.68 | 0.06 | 0.1751 | Lump/mass/lymph node | YELLOW |
|  | Mass of neck (4100606) | 2.86 | 0.03 | 0.2388 | Lump/mass/lymph node | ORANGE |
|  | Mass of skin (4183953) | 1.87 | 0.17 | 0.1698 | Skin Lesion | YELLOW |
|  | Mouth symptoms (4060063) | 3 | 0.06 | 0.2413 | Ulceration | ORANGE |
|  | Neck swelling (4200520) | 0.5 | 0 | 0.1003 | Lump/mass/lymph node | YELLOW |
|  | Pain in throat (259153) | 1.62 | 0.25 | 0.1427 | Respiratory | YELLOW |
|  | Pain of ear (4103476) | 2.61 | 0.69 | 0.1512 | Respiratory | YELLOW |
|  | Painful mouth (4010025) | 1.46 | 0.08 | 0.1584 | Ulceration | YELLOW |
|  | Skin lesion (4316083) | 3.55 | 1.65 | 0.1196 | Skin Lesion | YELLOW |
|  | Sore throat (4147326) | 5.06 | 1.13 | 0.2284 | Respiratory | ORANGE |
|  | Squamous cell carcinoma of skin (4111921) | 0.99 | 0.11 | 0.1192 | Skin Lesion | YELLOW |
|  | Swelling (4275722) | 2.15 | 0.5 | 0.1447 | Non-specific | YELLOW |
|  | Ulcer of mouth (4094986) | 6.3 | 0.11 | 0.357 | Ulceration | ORANGE |
|  | Ulcer on tongue (4278836) | 1.93 | 0 | 0.1984 | Ulceration | YELLOW |
| Liver | 10g monofilament sensation L foot abnormal (4214578) | 1.89 | 0.47 | 0.1318 | Other clinical findings | YELLOW |
|  | 10g monofilament sensation L foot normal (4215145) | 9.02 | 3.27 | 0.2412 | Other clinical findings | ORANGE |
|  | 10g monofilament sensation R foot abnormal (4215144) | 2.01 | 0.69 | 0.1146 | Other clinical findings | YELLOW |
|  | 10g monofilament sensation R foot normal (4214579) | 8.98 | 3.08 | 0.2498 | Other clinical findings | ORANGE |
|  | Abdominal pain (200219) | 8.31 | 1.56 | 0.3155 | Lower abdominal | ORANGE |
|  | Alcoholic cirrhosis (196463) | 1.36 | 0 | 0.1661 | Upper abdominal | YELLOW |
|  | Alcoholic liver damage (201612) | 0.61 | 0.02 | 0.1054 | Upper abdominal | YELLOW |
|  | All pulses present in left foot (37312546) | 5.96 | 1.76 | 0.2193 | Other clinical findings | ORANGE |
|  | All pulses present in left lower limb (37312545) | 7.3 | 3.22 | 0.1835 | Other clinical findings | YELLOW |
|  | All pulses present in right foot (37312547) | 5.94 | 1.8 | 0.2159 | Other clinical findings | ORANGE |
|  | All pulses present in right lower limb (37312544) | 7.28 | 3.16 | 0.186 | Other clinical findings | YELLOW |
|  | Anemia (439777) | 1.42 | 0.18 | 0.1395 | Non-specific | YELLOW |
|  | Ascites (200528) | 2.25 | 0 | 0.2146 | Upper abdominal | ORANGE |
|  | Backache (134736) | 3.65 | 1.72 | 0.1196 | Musculoskeletal | YELLOW |
|  | Blood pressure finding (4214962) | 73.55 | 57.74 | 0.3376 | Other clinical findings | ORANGE |
|  | Cellulitis (435613) | 2.31 | 0.85 | 0.1173 | Skin Lesion | YELLOW |
|  | Chest pain (77670) | 4.3 | 1.58 | 0.1615 | Respiratory | YELLOW |
|  | Cirrhosis of liver (4064161) | 4.77 | 0.04 | 0.3125 | Upper abdominal | ORANGE |
|  | Constipation (75860) | 3.33 | 1.48 | 0.121 | Lower abdominal | YELLOW |
|  | Cough (254761) | 9.73 | 6.43 | 0.1213 | Respiratory | YELLOW |
|  | Diabetes mellitus (201820) | 1.34 | 0.37 | 0.1055 | Upper abdominal | YELLOW |
|  | Diarrhea symptom (4145808) | 2.64 | 0.87 | 0.1351 | Lower abdominal | YELLOW |
|  | Disease of liver (194984) | 0.79 | 0 | 0.1262 | Upper abdominal | YELLOW |
|  | Duodenitis (433516) | 0.71 | 0.08 | 0.1006 | Upper abdominal | YELLOW |
|  | Dyspnea (312437) | 4.48 | 2.07 | 0.1357 | Respiratory | YELLOW |
|  | Edema (433595) | 2.17 | 0.79 | 0.1145 | Non-specific | YELLOW |
|  | Epigastric pain (197381) | 1.91 | 0.32 | 0.1519 | Upper abdominal | YELLOW |
|  | Esophageal varices (24966) | 2.11 | 0.02 | 0.2047 | Upper abdominal | ORANGE |
|  | Finding of foot region (4022924) | 3.93 | 1.68 | 0.1366 | Other clinical findings | YELLOW |
|  | Finding of pulse rate (4103199) | 17.48 | 11.09 | 0.1834 | Other clinical findings | YELLOW |
|  | Gastritis (201340) | 1.76 | 0.22 | 0.156 | Upper abdominal | YELLOW |
|  | Gastrointestinal hemorrhage (192671) | 0.63 | 0.04 | 0.1022 | Lower abdominal | YELLOW |
|  | Hemochromatosis (4163735) | 0.63 | 0.02 | 0.1073 | Non-specific | YELLOW |
|  | Hemorrhoids (195562) | 1.26 | 0.34 | 0.1034 | Lower abdominal | YELLOW |
|  | Hiatal hernia (4224004) | 1.28 | 0.37 | 0.1007 | Upper abdominal | YELLOW |
|  | Hypochondrial pain (4156030) | 0.67 | 0.02 | 0.111 | Upper abdominal | YELLOW |
|  | Iron deficiency anemia (436659) | 1.81 | 0.43 | 0.1314 | Non-specific | YELLOW |
|  | Jaundice (137977) | 1.26 | 0 | 0.1598 | Upper abdominal | YELLOW |
|  | Large liver (197676) | 0.77 | 0 | 0.1246 | Upper abdominal | YELLOW |
|  | Lower respiratory tract infection (4175297) | 4.52 | 2.45 | 0.113 | Respiratory | YELLOW |
|  | Nausea (31967) | 1.62 | 0.34 | 0.1302 | Upper abdominal | YELLOW |
|  | Nonproliferative retinopathy of left eye due to diabetes mellitus (37311254) | 1.81 | 0.53 | 0.1192 | Non-specific | YELLOW |
|  | Normal pulse in left dorsalis pedis artery (37311332) | 1.7 | 0.53 | 0.1116 | Other clinical findings | YELLOW |
|  | Normal pulse in right dorsalis pedis artery (37311331) | 1.66 | 0.51 | 0.1112 | Other clinical findings | YELLOW |
|  | Normal tactile sensation (37311136) | 2.58 | 1.03 | 0.1166 | Other clinical findings | YELLOW |
|  | Obstructive hyperbilirubinemia (4242900) | 0.59 | 0 | 0.1089 | Upper abdominal | YELLOW |
|  | Portal hypertension (192680) | 1.38 | 0.02 | 0.1637 | Upper abdominal | YELLOW |
|  | Pruritic disorder (136184) | 1.32 | 0.34 | 0.1082 | Non-specific | YELLOW |
|  | Type 2 diabetes mellitus (201826) | 2.74 | 1.26 | 0.1059 | Upper abdominal | YELLOW |
|  | Ulcer of duodenum (4198381) | 0.63 | 0 | 0.1126 | Upper abdominal | YELLOW |
|  | Venous varices (312349) | 0.83 | 0.02 | 0.1248 | Non-specific | YELLOW |
|  | Vomiting (441408) | 1.48 | 0.3 | 0.1259 | Upper abdominal | YELLOW |
| Lung | Acute exacerbation of chronic obstructive pulmonary disease (257004) | 3.4 | 0.61 | 0.2 | Respiratory | YELLOW |
|  | Acute lower respiratory tract infection (4307774) | 5.67 | 1.74 | 0.2092 | Respiratory | ORANGE |
|  | Backache (134736) | 4.4 | 2.09 | 0.1306 | Musculoskeletal | YELLOW |
|  | Blood pressure finding (4214962) | 70.49 | 60.94 | 0.2022 | Other clinical findings | ORANGE |
|  | Chest pain (77670) | 6.73 | 2.15 | 0.2237 | Respiratory | ORANGE |
|  | Chronic obstructive pulmonary disease (255573) | 4.53 | 0.77 | 0.2357 | Respiratory | ORANGE |
|  | Constipation (75860) | 3.63 | 1.9 | 0.1057 | Lower abdominal | YELLOW |
|  | Cough (254761) | 20.83 | 7.2 | 0.4004 | Respiratory | RED |
|  | Dyspnea (312437) | 10.45 | 2.72 | 0.3155 | Respiratory | ORANGE |
|  | Finding of pulse rate (4103199) | 16.57 | 11.24 | 0.1545 | Other clinical findings | YELLOW |
|  | Hemoptysis (261687) | 4.06 | 0.2 | 0.2698 | Respiratory | ORANGE |
|  | Infective pneumonia (443410) | 0.88 | 0.12 | 0.1079 | Respiratory | YELLOW |
|  | Lower respiratory tract infection (4175297) | 8.48 | 2.66 | 0.2558 | Respiratory | ORANGE |
|  | Pain (4329041) | 2.94 | 1.39 | 0.1067 | Non-specific | YELLOW |
|  | Pleural effusion (254061) | 1.26 | 0.08 | 0.145 | Respiratory | YELLOW |
|  | Productive cough (4102774) | 3.1 | 1.05 | 0.1442 | Respiratory | YELLOW |
| Nasopharynx | 10g monofilament sensation L foot normal (4215145) | 2.66 | 1.21 | 0.1054 | Other clinical findings | YELLOW |
|  | Acute secretory otitis media (374948) | 0.72 | 0 | 0.1204 | Non-specific | YELLOW |
|  | Acute sinusitis (260123) | 4.35 | 0.48 | 0.2541 | Respiratory | ORANGE |
|  | Acute tonsillitis (24660) | 1.69 | 0 | 0.1854 | Respiratory | YELLOW |
|  | Alcohol dependence (435243) | 1.21 | 0.24 | 0.1145 | Non-specific | YELLOW |
|  | Allergic reaction to drug (4166257) | 4.83 | 1.45 | 0.1947 | Non-specific | YELLOW |
|  | Allergic rhinitis (257007) | 0.72 | 0 | 0.1204 | Respiratory | YELLOW |
|  | Anxiety (441542) | 1.45 | 0.24 | 0.1325 | Non-specific | YELLOW |
|  | Bleeding from nose (4096682) | 5.56 | 0.97 | 0.2605 | Respiratory | ORANGE |
|  | Blood pressure finding (4214962) | 55.8 | 50.24 | 0.1116 | Other clinical findings | YELLOW |
|  | Candidiasis of mouth and esophagus (4142836) | 0.72 | 0 | 0.1204 | Upper abdominal | YELLOW |
|  | Cervical lymphadenopathy (4133459) | 2.42 | 0 | 0.2227 | Lump/mass/lymph node | ORANGE |
|  | Chronic catarrhal rhinitis (4145495) | 0.97 | 0 | 0.14 | Respiratory | YELLOW |
|  | Chronic rhinitis (259848) | 1.45 | 0.24 | 0.1325 | Respiratory | YELLOW |
|  | Chronic sinusitis (257012) | 1.45 | 0 | 0.1715 | Respiratory | YELLOW |
|  | Community acquired pneumonia (4293463) | 0.72 | 0 | 0.1204 | Respiratory | YELLOW |
|  | Cough (254761) | 9.42 | 3.38 | 0.2487 | Respiratory | ORANGE |
|  | Diplopia (373474) | 0.97 | 0 | 0.14 | Central nervous system | YELLOW |
|  | Discomfort in mouth (4150136) | 0.72 | 0 | 0.1204 | Ulceration | YELLOW |
|  | Dysfunction of eustachian tube (381877) | 2.66 | 0.48 | 0.176 | Respiratory | YELLOW |
|  | Dysphagia (31317) | 2.9 | 0.24 | 0.2152 | Upper abdominal | ORANGE |
|  | Epidermoid cyst of skin (137351) | 0.72 | 0 | 0.1204 | Skin Lesion | YELLOW |
|  | Eruption (140214) | 3.38 | 0.48 | 0.212 | Skin Lesion | ORANGE |
|  | Essential hypertension (320128) | 1.93 | 0.72 | 0.106 | Non-specific | YELLOW |
|  | Feeling of lump in throat (4145818) | 0.72 | 0 | 0.1204 | Respiratory | YELLOW |
|  | Finding of head and neck region (255919) | 1.45 | 0 | 0.1715 | Lump/mass/lymph node | YELLOW |
|  | Finding of pharynx (4022571) | 0.72 | 0 | 0.1204 | Respiratory | YELLOW |
|  | Gastric reflux (4023183) | 0.97 | 0 | 0.14 | Upper abdominal | YELLOW |
|  | Gastritis (201340) | 0.97 | 0 | 0.14 | Upper abdominal | YELLOW |
|  | Headache (378253) | 7.25 | 0.97 | 0.3204 | Central nervous system | ORANGE |
|  | Hearing loss (377889) | 4.59 | 0.97 | 0.2215 | Respiratory | ORANGE |
|  | Hemoptysis (261687) | 1.45 | 0.24 | 0.1325 | Respiratory | YELLOW |
|  | Hyperlipidemia (432867) | 0.72 | 0 | 0.1204 | Non-specific | YELLOW |
|  | Infective otitis externa (381859) | 3.14 | 0.48 | 0.2005 | Respiratory | ORANGE |
|  | Inflammatory dermatosis (45766714) | 0.97 | 0 | 0.14 | Skin Lesion | YELLOW |
|  | Insomnia (436962) | 2.66 | 1.21 | 0.1054 | Non-specific | YELLOW |
|  | Iron deficiency anemia (436659) | 0.97 | 0 | 0.14 | Non-specific | YELLOW |
|  | Jaw pain (4168686) | 0.72 | 0 | 0.1204 | Non-specific | YELLOW |
|  | Lower respiratory tract infection (4175297) | 3.38 | 1.45 | 0.126 | Respiratory | YELLOW |
|  | Lymphadenopathy (315085) | 4.83 | 0 | 0.3186 | Lump/mass/lymph node | ORANGE |
|  | Mass of neck (4100606) | 8.45 | 0 | 0.4296 | Lump/mass/lymph node | RED |
|  | Mass of skin (4183953) | 4.35 | 0 | 0.3016 | Skin Lesion | ORANGE |
|  | Mouth symptoms (4060063) | 1.21 | 0 | 0.1565 | Ulceration | YELLOW |
|  | Nasal congestion (4195085) | 1.69 | 0 | 0.1854 | Respiratory | YELLOW |
|  | Nasal symptom (4091808) | 2.66 | 0.48 | 0.176 | Respiratory | YELLOW |
|  | Neck pain (24134) | 1.93 | 0.72 | 0.106 | Musculoskeletal | YELLOW |
|  | Neck swelling (4200520) | 0.97 | 0 | 0.14 | Lump/mass/lymph node | YELLOW |
|  | Non-suppurative otitis media with eustachian tube disorder (4111114) | 0.72 | 0 | 0.1204 | Respiratory | YELLOW |
|  | Normal pulse rate (4026687) | 0.72 | 0 | 0.1204 | Other clinical findings | YELLOW |
|  | Normal tactile sensation (37311136) | 1.21 | 0.24 | 0.1145 | Other clinical findings | YELLOW |
|  | Pain in face (4317968) | 0.72 | 0 | 0.1204 | Non-specific | YELLOW |
|  | Pain in throat (259153) | 1.69 | 0.24 | 0.1487 | Respiratory | YELLOW |
|  | Pain of ear (4103476) | 5.31 | 0.48 | 0.2911 | Respiratory | ORANGE |
|  | Parotid swelling (4152183) | 0.97 | 0 | 0.14 | Lump/mass/lymph node | YELLOW |
|  | Polyp of nasal cavity and/or nasal sinus (42537251) | 1.21 | 0 | 0.1565 | Respiratory | YELLOW |
|  | Posterior rhinorrhea (257683) | 0.97 | 0 | 0.14 | Respiratory | YELLOW |
|  | Purulent otitis media (376712) | 0.72 | 0 | 0.1204 | Respiratory | YELLOW |
|  | Sensation of blocked ear (4011332) | 3.38 | 0.97 | 0.1658 | Respiratory | YELLOW |
|  | Sinus headache (4172302) | 1.93 | 0 | 0.1984 | Respiratory | YELLOW |
|  | Sinusitis (4283893) | 1.21 | 0 | 0.1565 | Respiratory | YELLOW |
|  | Sore throat (4147326) | 7.49 | 0.97 | 0.3283 | Respiratory | ORANGE |
|  | Swallowing symptoms (4149859) | 0.72 | 0 | 0.1204 | Upper abdominal | YELLOW |
|  | Swelling (4275722) | 3.38 | 0 | 0.2645 | Non-specific | ORANGE |
|  | Wax in ear canal (4155902) | 3.14 | 1.45 | 0.113 | Respiratory | YELLOW |
| Oropharynx | Acute pharyngitis (25297) | 1.76 | 0.08 | 0.1766 | Respiratory | YELLOW |
|  | Acute tonsillitis (24660) | 3.52 | 0 | 0.2701 | Respiratory | ORANGE |
|  | Blood pressure finding (4214962) | 55.34 | 48.83 | 0.1306 | Other clinical findings | YELLOW |
|  | Cervical lymphadenopathy (4133459) | 1.68 | 0 | 0.1849 | Lump/mass/lymph node | YELLOW |
|  | Disorder of tongue (432941) | 0.77 | 0.04 | 0.1151 | Ulceration | YELLOW |
|  | Dysphagia (31317) | 2.99 | 0.23 | 0.2206 | Upper abdominal | ORANGE |
|  | Ear finding (4082416) | 0.65 | 0.04 | 0.1042 | Respiratory | YELLOW |
|  | Feeling of lump in throat (4145818) | 1.38 | 0 | 0.1673 | Respiratory | YELLOW |
|  | Finding of head and neck region (255919) | 1.34 | 0.04 | 0.1575 | Lump/mass/lymph node | YELLOW |
|  | Finding of lymph node (4200648) | 0.5 | 0 | 0.1003 | Lump/mass/lymph node | YELLOW |
|  | Finding of pharynx (4022571) | 0.84 | 0 | 0.1302 | Respiratory | YELLOW |
|  | Hemoptysis (261687) | 0.84 | 0.08 | 0.1125 | Respiratory | YELLOW |
|  | Jaw pain (4168686) | 0.5 | 0 | 0.1003 | Non-specific | YELLOW |
|  | Lymphadenopathy (315085) | 5.09 | 0.08 | 0.3197 | Lump/mass/lymph node | ORANGE |
|  | Mass of neck (4100606) | 11.52 | 0.04 | 0.5075 | Lump/mass/lymph node | RED |
|  | Mass of skin (4183953) | 4.06 | 0.31 | 0.2586 | Skin Lesion | ORANGE |
|  | Mouth symptoms (4060063) | 1.03 | 0.15 | 0.1151 | Ulceration | YELLOW |
|  | Neck swelling (4200520) | 1.26 | 0 | 0.1598 | Lump/mass/lymph node | YELLOW |
|  | Pain in throat (259153) | 3.75 | 0.08 | 0.2702 | Respiratory | ORANGE |
|  | Pain of ear (4103476) | 4.78 | 0.65 | 0.2562 | Respiratory | ORANGE |
|  | Painful mouth (4010025) | 0.69 | 0.04 | 0.1079 | Ulceration | YELLOW |
|  | Parotid swelling (4152183) | 0.5 | 0 | 0.1003 | Lump/mass/lymph node | YELLOW |
|  | Sore throat (4147326) | 14.43 | 0.84 | 0.5294 | Respiratory | RED |
|  | Squamous cell carcinoma of skin (4111921) | 0.8 | 0.08 | 0.1089 | Skin Lesion | YELLOW |
|  | Swelling (4275722) | 4.36 | 0.27 | 0.2745 | Non-specific | ORANGE |
|  | Ulcer of mouth (4094986) | 1.38 | 0.15 | 0.1415 | Ulceration | YELLOW |
| Ovary | Abdominal bloating (4023572) | 2.82 | 0.29 | 0.2056 | Lower abdominal | ORANGE |
|  | Abdominal discomfort (4183041) | 1.83 | 0.18 | 0.166 | Lower abdominal | YELLOW |
|  | Abdominal distension symptom (4012079) | 2.35 | 0.08 | 0.2083 | Lower abdominal | ORANGE |
|  | Abdominal mass (192438) | 2.59 | 0.02 | 0.2279 | Lump/mass/lymph node | ORANGE |
|  | Abdominal pain (200219) | 17.03 | 2.34 | 0.5128 | Lower abdominal | RED |
|  | Ascites (200528) | 1.74 | 0 | 0.1882 | Upper abdominal | YELLOW |
|  | Blood pressure finding (4214962) | 60.94 | 55.74 | 0.1056 | Other clinical findings | YELLOW |
|  | Constipation (75860) | 6.05 | 1.56 | 0.2363 | Lower abdominal | ORANGE |
|  | Cyst of ovary (197610) | 5.01 | 0.09 | 0.316 | Female specific/breast | ORANGE |
|  | Deep venous thrombosis (4133004) | 1.1 | 0.08 | 0.1335 | Non-specific | YELLOW |
|  | Epigastric pain (197381) | 1.74 | 0.51 | 0.1168 | Upper abdominal | YELLOW |
|  | Finding of abdominopelvic segment of trunk (37311678) | 2.15 | 0.02 | 0.2067 | Lower abdominal | ORANGE |
|  | Irritable bowel syndrome (75576) | 1.74 | 0.49 | 0.1193 | Lower abdominal | YELLOW |
|  | Nausea (31967) | 1.97 | 0.49 | 0.1346 | Upper abdominal | YELLOW |
|  | Pain in pelvis (4147829) | 1.14 | 0.12 | 0.1292 | Lower abdominal | YELLOW |
|  | Pelvic mass (201131) | 2.81 | 0 | 0.2405 | Lump/mass/lymph node | ORANGE |
|  | Pleural effusion (254061) | 1.07 | 0.07 | 0.1331 | Respiratory | YELLOW |
|  | Swollen abdomen (442597) | 0.6 | 0.03 | 0.1019 | Lower abdominal | YELLOW |
|  | Vomiting (441408) | 1.99 | 0.5 | 0.1347 | Upper abdominal | YELLOW |
| Pancreas | 10g monofilament sensation L foot normal (4215145) | 5.88 | 2.98 | 0.1413 | Other clinical findings | YELLOW |
|  | 10g monofilament sensation R foot normal (4214579) | 6 | 2.9 | 0.1508 | Other clinical findings | YELLOW |
|  | Abdomen examined - NAD (4060865) | 2 | 0.37 | 0.1511 | Other clinical findings | YELLOW |
|  | Abdominal bloating (4023572) | 0.97 | 0.14 | 0.1119 | Lower abdominal | YELLOW |
|  | Abdominal discomfort (4183041) | 1.75 | 0.15 | 0.1655 | Lower abdominal | YELLOW |
|  | Abdominal pain (200219) | 17.37 | 2.19 | 0.5286 | Lower abdominal | RED |
|  | Acute pancreatitis (199074) | 0.97 | 0.04 | 0.1315 | Upper abdominal | YELLOW |
|  | All pulses present in left foot (37312546) | 3.22 | 1.65 | 0.102 | Other clinical findings | YELLOW |
|  | All pulses present in left lower limb (37312545) | 5.67 | 3.08 | 0.1269 | Other clinical findings | YELLOW |
|  | All pulses present in right foot (37312547) | 3.2 | 1.66 | 0.1001 | Other clinical findings | YELLOW |
|  | All pulses present in right lower limb (37312544) | 5.69 | 3.06 | 0.1288 | Other clinical findings | YELLOW |
|  | Backache (134736) | 5.34 | 2.21 | 0.1648 | Musculoskeletal | YELLOW |
|  | Blood pressure finding (4214962) | 71.69 | 60.42 | 0.2397 | Other clinical findings | ORANGE |
|  | Carcinoma in situ of pancreas (4245553) | 0.65 | 0 | 0.1144 | Upper abdominal | YELLOW |
|  | Chest pain (77670) | 3.87 | 1.94 | 0.1151 | Respiratory | YELLOW |
|  | Chronic pancreatitis (195596) | 0.56 | 0 | 0.1061 | Upper abdominal | YELLOW |
|  | Constipation (75860) | 6.77 | 2.02 | 0.2333 | Lower abdominal | ORANGE |
|  | Diabetes mellitus (201820) | 2.83 | 0.53 | 0.1797 | Upper abdominal | YELLOW |
|  | Diarrhea (196523) | 1.61 | 0.36 | 0.1268 | Lower abdominal | YELLOW |
|  | Diarrhea symptom (4145808) | 3.29 | 1.3 | 0.1332 | Lower abdominal | YELLOW |
|  | Epigastric pain (197381) | 6.14 | 0.36 | 0.3304 | Upper abdominal | ORANGE |
|  | Gallstone (196456) | 1.23 | 0.26 | 0.113 | Upper abdominal | YELLOW |
|  | Gastritis (201340) | 1.43 | 0.27 | 0.1266 | Upper abdominal | YELLOW |
|  | Gastro-esophageal reflux disease with esophagitis (30437) | 1.04 | 0.21 | 0.1055 | Upper abdominal | YELLOW |
|  | Gastroduodenitis (195306) | 0.98 | 0.09 | 0.1222 | Upper abdominal | YELLOW |
|  | Hiatal hernia (4224004) | 1.76 | 0.47 | 0.1231 | Upper abdominal | YELLOW |
|  | Hypoglycemia (24609) | 0.78 | 0.11 | 0.1008 | Upper abdominal | YELLOW |
|  | Jaundice (137977) | 5.78 | 0.04 | 0.3466 | Upper abdominal | ORANGE |
|  | Loin pain (4152353) | 0.95 | 0.18 | 0.1029 | Genitourinary | YELLOW |
|  | Low back pain (194133) | 3.92 | 2.14 | 0.104 | Musculoskeletal | YELLOW |
|  | Nausea (31967) | 2.12 | 0.49 | 0.144 | Upper abdominal | YELLOW |
|  | Obstructive hyperbilirubinemia (4242900) | 3.51 | 0.02 | 0.2674 | Upper abdominal | ORANGE |
|  | Pain (4329041) | 3.21 | 1.29 | 0.1297 | Non-specific | YELLOW |
|  | Pruritic disorder (136184) | 1.82 | 0.46 | 0.1284 | Non-specific | YELLOW |
|  | Type 2 diabetes mellitus (201826) | 6.2 | 1.3 | 0.2601 | Upper abdominal | ORANGE |
|  | Vomiting (441408) | 2.48 | 0.43 | 0.1718 | Upper abdominal | YELLOW |
| Prostate | Benign prostatic hyperplasia (198803) | 2.64 | 0.58 | 0.1642 | Genitourinary | YELLOW |
|  | Blood pressure finding (4214962) | 70.97 | 60.78 | 0.2162 | Other clinical findings | ORANGE |
|  | Carcinoma in situ of prostate (200970) | 0.72 | 0.03 | 0.1131 | Genitourinary | YELLOW |
|  | Dysuria (197684) | 2.03 | 0.52 | 0.1349 | Genitourinary | YELLOW |
|  | Low back pain (194133) | 3.71 | 1.86 | 0.1126 | Musculoskeletal | YELLOW |
|  | Nocturia (40304526) | 4.23 | 0.71 | 0.2283 | Genitourinary | ORANGE |
|  | Prostatism (4016155) | 7.16 | 0.84 | 0.3268 | Genitourinary | ORANGE |
|  | Retention of urine (192450) | 2.86 | 0.37 | 0.1985 | Genitourinary | YELLOW |
|  | Urinary tract infectious disease (81902) | 2.87 | 0.97 | 0.1388 | Genitourinary | YELLOW |
| Rectum | Abdomen examined - NAD (4060865) | 1.53 | 0.38 | 0.1185 | Other clinical findings | YELLOW |
|  | Abdominal pain (200219) | 4.17 | 1.92 | 0.1312 | Lower abdominal | YELLOW |
|  | Constipation (75860) | 6.04 | 1.77 | 0.2218 | Lower abdominal | ORANGE |
|  | Diarrhea (196523) | 2.26 | 0.45 | 0.157 | Lower abdominal | YELLOW |
|  | Diarrhea symptom (4145808) | 5.53 | 1.09 | 0.2501 | Lower abdominal | ORANGE |
|  | Diverticular disease (4266809) | 1.68 | 0.54 | 0.109 | Lower abdominal | YELLOW |
|  | External hemorrhoids without complication (199839) | 0.92 | 0.08 | 0.1193 | Lower abdominal | YELLOW |
|  | Hematochezia (443530) | 0.62 | 0.02 | 0.1064 | Lower abdominal | YELLOW |
|  | Hemorrhoids (195562) | 3.56 | 0.48 | 0.2203 | Lower abdominal | ORANGE |
|  | Incontinence of feces (197675) | 0.71 | 0.06 | 0.1051 | Lower abdominal | YELLOW |
|  | Internal hemorrhoids grade I (36715916) | 0.8 | 0.08 | 0.1089 | Lower abdominal | YELLOW |
|  | Iron deficiency anemia (436659) | 1.77 | 0.43 | 0.1287 | Non-specific | YELLOW |
|  | Loose stool (4161687) | 0.69 | 0.05 | 0.1056 | Lower abdominal | YELLOW |
|  | Painful rectal bleeding (4189790) | 0.88 | 0.04 | 0.1244 | Lower abdominal | YELLOW |
|  | Polyp of colon (4285898) | 1.24 | 0.11 | 0.1383 | Lower abdominal | YELLOW |
|  | Rectal hemorrhage (4026112) | 15.33 | 0.5 | 0.5713 | Lower abdominal | RED |
|  | Rectal polyp (4216644) | 2.09 | 0.04 | 0.2007 | Lower abdominal | ORANGE |
| Salivary gland | Actinic keratosis (138825) | 2.33 | 0.74 | 0.1296 | Skin Lesion | YELLOW |
|  | Adenolymphoma (4307552) | 0.74 | 0 | 0.1221 | Lump/mass/lymph node | YELLOW |
|  | Basal cell carcinoma of skin (4112752) | 1.97 | 0.25 | 0.1647 | Skin Lesion | YELLOW |
|  | Bell's palsy (374923) | 3.07 | 0 | 0.2517 | Central nervous system | ORANGE |
|  | Blood pressure finding (4214962) | 59.34 | 52.46 | 0.1389 | Other clinical findings | YELLOW |
|  | Cervical lymphadenopathy (4133459) | 0.86 | 0 | 0.1317 | Lump/mass/lymph node | YELLOW |
|  | Diarrhea symptom (4145808) | 1.35 | 0.25 | 0.1237 | Lower abdominal | YELLOW |
|  | Disease (4274025) | 1.23 | 0.25 | 0.1145 | Non-specific | YELLOW |
|  | Ear, nose and throat finding (4178545) | 0.86 | 0 | 0.1317 | Respiratory | YELLOW |
|  | Eruption (140214) | 3.56 | 1.6 | 0.1239 | Skin Lesion | YELLOW |
|  | Essential hypertension (320128) | 2.58 | 0.74 | 0.1444 | Non-specific | YELLOW |
|  | Eye symptom (4205680) | 1.23 | 0.12 | 0.1359 | Non-specific | YELLOW |
|  | Facial swelling (4133326) | 0.98 | 0.12 | 0.1165 | Respiratory | YELLOW |
|  | Finding of head and neck region (255919) | 0.74 | 0 | 0.1221 | Lump/mass/lymph node | YELLOW |
|  | Finding of lymph node (4200648) | 0.86 | 0 | 0.1317 | Lump/mass/lymph node | YELLOW |
|  | Finding of neck region (4184252) | 0.61 | 0 | 0.1108 | Lump/mass/lymph node | YELLOW |
|  | Headache (378253) | 2.33 | 0.86 | 0.1175 | Central nervous system | YELLOW |
|  | Jaw pain (4168686) | 1.23 | 0.12 | 0.1359 | Non-specific | YELLOW |
|  | Lymphadenopathy (315085) | 5.04 | 0 | 0.3258 | Lump/mass/lymph node | ORANGE |
|  | Mass of neck (4100606) | 8.97 | 0.12 | 0.4348 | Lump/mass/lymph node | RED |
|  | Mass of parotid gland (4202556) | 4.67 | 0 | 0.313 | Lump/mass/lymph node | ORANGE |
|  | Mass of skin (4183953) | 8.72 | 0 | 0.4371 | Skin Lesion | RED |
|  | Neoplasm of uncertain behavior of parotid gland (4313657) | 0.74 | 0 | 0.1221 | Lump/mass/lymph node | YELLOW |
|  | Pain in face (4317968) | 1.11 | 0 | 0.1498 | Non-specific | YELLOW |
|  | Pain of ear (4103476) | 3.69 | 0.86 | 0.1907 | Respiratory | YELLOW |
|  | Parotid swelling (4152183) | 8.6 | 0.12 | 0.4245 | Lump/mass/lymph node | RED |
|  | Parotitis (4032796) | 0.98 | 0 | 0.1407 | Non-specific | YELLOW |
|  | Peripheral edema (4158343) | 1.11 | 0.25 | 0.1048 | Non-specific | YELLOW |
|  | Skin lesion (4316083) | 2.83 | 0.98 | 0.1356 | Skin Lesion | YELLOW |
|  | Sore throat (4147326) | 2.09 | 0.86 | 0.1022 | Respiratory | YELLOW |
|  | Squamous cell carcinoma of skin (4111921) | 1.11 | 0 | 0.1498 | Skin Lesion | YELLOW |
|  | Submandibular salivary gland swelling (4152184) | 0.98 | 0 | 0.1407 | Lump/mass/lymph node | YELLOW |
|  | Swelling (4275722) | 10.81 | 0.74 | 0.4421 | Non-specific | RED |
| Stomach | Abdomen examined - NAD (4060865) | 2.1 | 0.63 | 0.1269 | Other clinical findings | YELLOW |
|  | Abdominal discomfort (4183041) | 1 | 0.17 | 0.109 | Lower abdominal | YELLOW |
|  | Abdominal pain (200219) | 9.36 | 2.23 | 0.3088 | Lower abdominal | ORANGE |
|  | Acid reflux (44783954) | 1.03 | 0.16 | 0.1133 | Upper abdominal | YELLOW |
|  | Anemia (439777) | 3.14 | 0.34 | 0.2154 | Non-specific | ORANGE |
|  | Blood pressure finding (4214962) | 72.83 | 63.65 | 0.1982 | Other clinical findings | YELLOW |
|  | Chest pain (77670) | 5.7 | 2.32 | 0.1729 | Respiratory | YELLOW |
|  | Constipation (75860) | 5.89 | 2.01 | 0.2002 | Lower abdominal | ORANGE |
|  | Dysphagia (31317) | 3.73 | 0.47 | 0.2288 | Upper abdominal | ORANGE |
|  | Dyspnea (312437) | 5.43 | 3.06 | 0.1178 | Respiratory | YELLOW |
|  | Epigastric pain (197381) | 5.68 | 0.53 | 0.3002 | Upper abdominal | ORANGE |
|  | Finding of pulse rate (4103199) | 15.14 | 10.96 | 0.1243 | Other clinical findings | YELLOW |
|  | Gastric ulcer (4265600) | 3.87 | 0.13 | 0.2696 | Upper abdominal | ORANGE |
|  | Gastritis (201340) | 1.41 | 0.3 | 0.1208 | Upper abdominal | YELLOW |
|  | Gastro-esophageal reflux disease with esophagitis (30437) | 1.69 | 0.43 | 0.1233 | Upper abdominal | YELLOW |
|  | Gastroesophageal reflux disease (318800) | 1.46 | 0.3 | 0.1244 | Upper abdominal | YELLOW |
|  | Gastrointestinal hemorrhage (192671) | 0.87 | 0.07 | 0.1172 | Lower abdominal | YELLOW |
|  | Heartburn (23325) | 1.5 | 0.25 | 0.1345 | Upper abdominal | YELLOW |
|  | Hematemesis (26727) | 0.85 | 0.1 | 0.1092 | Upper abdominal | YELLOW |
|  | Hiatal hernia (4224004) | 1.5 | 0.47 | 0.1044 | Upper abdominal | YELLOW |
|  | Iron deficiency anemia (436659) | 6.28 | 0.47 | 0.326 | Non-specific | ORANGE |
|  | Melena (4103703) | 1.09 | 0.09 | 0.1309 | Lower abdominal | YELLOW |
|  | Microcytic hypochromic anemia (4262948) | 2.5 | 0.1 | 0.2131 | Non-specific | ORANGE |
|  | Nausea (31967) | 2.46 | 0.59 | 0.153 | Upper abdominal | YELLOW |
|  | Vomiting (441408) | 3.97 | 0.57 | 0.2298 | Upper abdominal | ORANGE |
| Thyroid | Acquired hypothyroidism (138384) | 1.51 | 0.17 | 0.1472 | Non-specific | YELLOW |
|  | Blood pressure finding (4214962) | 55.71 | 44.09 | 0.234 | Other clinical findings | ORANGE |
|  | Carcinoma in situ of thyroid gland (4241917) | 0.96 | 0 | 0.1392 | Lump/mass/lymph node | YELLOW |
|  | Cyst of thyroid (138113) | 2.43 | 0 | 0.2232 | Lump/mass/lymph node | ORANGE |
|  | Disorder of thyroid gland (141253) | 1.95 | 0 | 0.1994 | Non-specific | YELLOW |
|  | Dysphagia (31317) | 1.27 | 0.27 | 0.1146 | Upper abdominal | YELLOW |
|  | Essential hypertension (320128) | 2.02 | 0.82 | 0.1016 | Non-specific | YELLOW |
|  | Feeling of lump in throat (4145818) | 0.89 | 0.14 | 0.1049 | Respiratory | YELLOW |
|  | Finding of head and neck region (255919) | 2.37 | 0 | 0.2203 | Lump/mass/lymph node | ORANGE |
|  | Finding of neck region (4184252) | 0.89 | 0 | 0.134 | Lump/mass/lymph node | YELLOW |
|  | Finding of thyroid gland (4201376) | 2.43 | 0 | 0.2232 | Lump/mass/lymph node | ORANGE |
|  | Goiter (135772) | 2.81 | 0 | 0.2405 | Lump/mass/lymph node | ORANGE |
|  | Graves' disease (4232076) | 0.62 | 0 | 0.1117 | Non-specific | YELLOW |
|  | Hyperparathyroidism (133729) | 0.51 | 0 | 0.1013 | Non-specific | YELLOW |
|  | Hyperthyroidism (4142479) | 0.96 | 0 | 0.1392 | Non-specific | YELLOW |
|  | Hypothyroidism (140673) | 2.71 | 0.48 | 0.1787 | Non-specific | YELLOW |
|  | Lymphadenopathy (315085) | 1.65 | 0.14 | 0.1608 | Lump/mass/lymph node | YELLOW |
|  | Mass of neck (4100606) | 12.79 | 0.1 | 0.535 | Lump/mass/lymph node | RED |
|  | Mass of skin (4183953) | 5.93 | 0.24 | 0.3336 | Skin Lesion | ORANGE |
|  | Neck pain (24134) | 2.78 | 0.99 | 0.1319 | Musculoskeletal | YELLOW |
|  | Neck swelling (4200520) | 2.06 | 0 | 0.2051 | Lump/mass/lymph node | ORANGE |
|  | Non-toxic multinodular goiter (136368) | 2.54 | 0 | 0.2283 | Lump/mass/lymph node | ORANGE |
|  | Non-toxic nodular goiter (136661) | 0.93 | 0 | 0.137 | Lump/mass/lymph node | YELLOW |
|  | Postoperative hypothyroidism (137820) | 0.51 | 0 | 0.1013 | Non-specific | YELLOW |
|  | Simple goiter (141825) | 6.79 | 0 | 0.3817 | Lump/mass/lymph node | ORANGE |
|  | Sore throat (4147326) | 4.53 | 1.68 | 0.1649 | Respiratory | YELLOW |
|  | Swelling (4275722) | 4.08 | 0.38 | 0.2526 | Non-specific | ORANGE |
|  | Thyroid nodule (4030041) | 9.56 | 0 | 0.4598 | Lump/mass/lymph node | RED |
|  | Thyrotoxicosis (138387) | 1.03 | 0.07 | 0.1301 | Non-specific | YELLOW |

*lung cancer includes cancer of lung, bronchus and trachea

### **Figure S1: Site-specific clinical concepts with absolute standardized mean differences (SMD) > 0.1 for lung cancer. Colours represent the magnitude of the SMD, with higher values indicating greater differences between cases and matched individuals.**

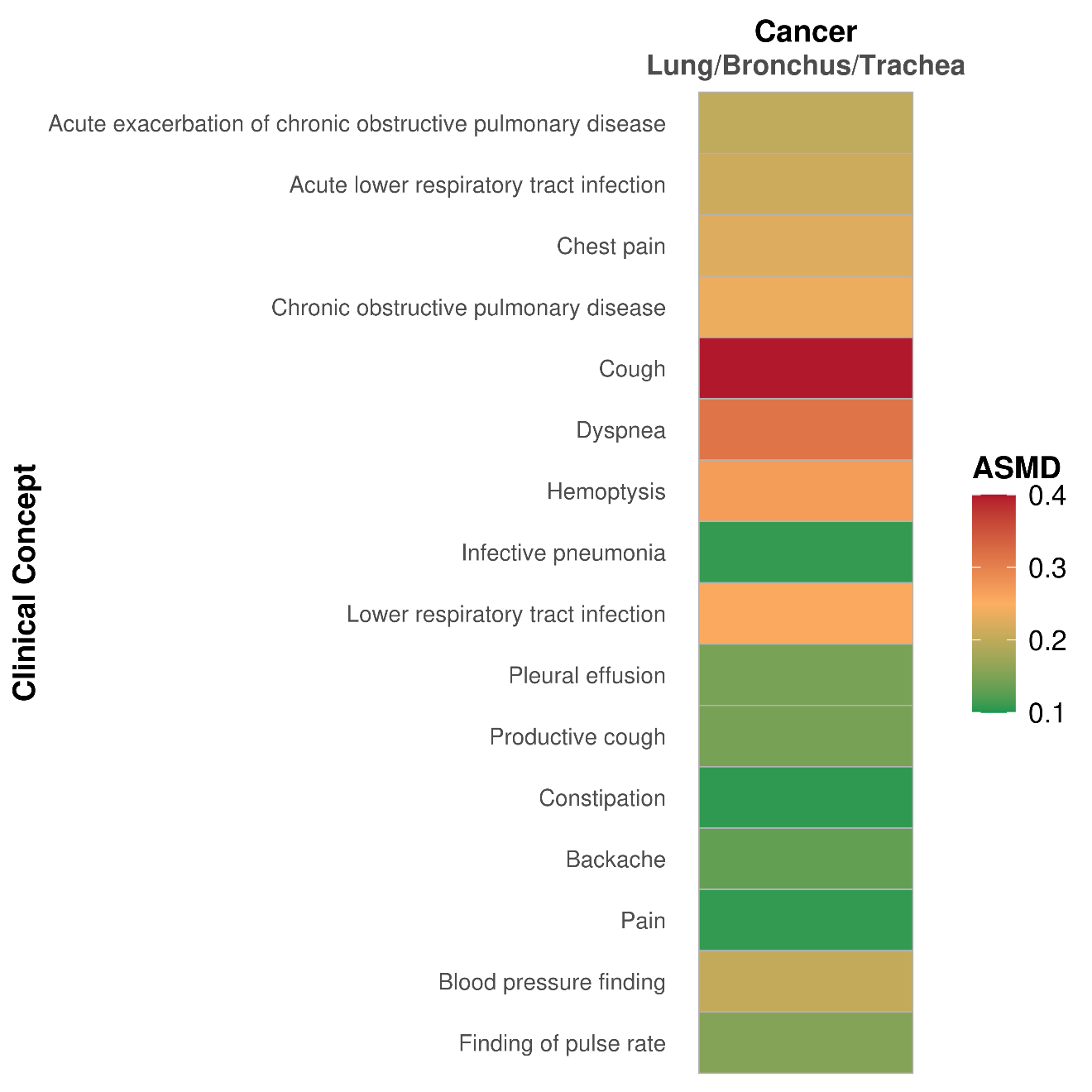

### **Figure S2: Site-specific clinical concepts with standardized mean differences (SMD) > 0.1 for genitourinary cancers. Colours represent the magnitude of the SMD, with higher values indicating greater differences between cases and matched individuals.**

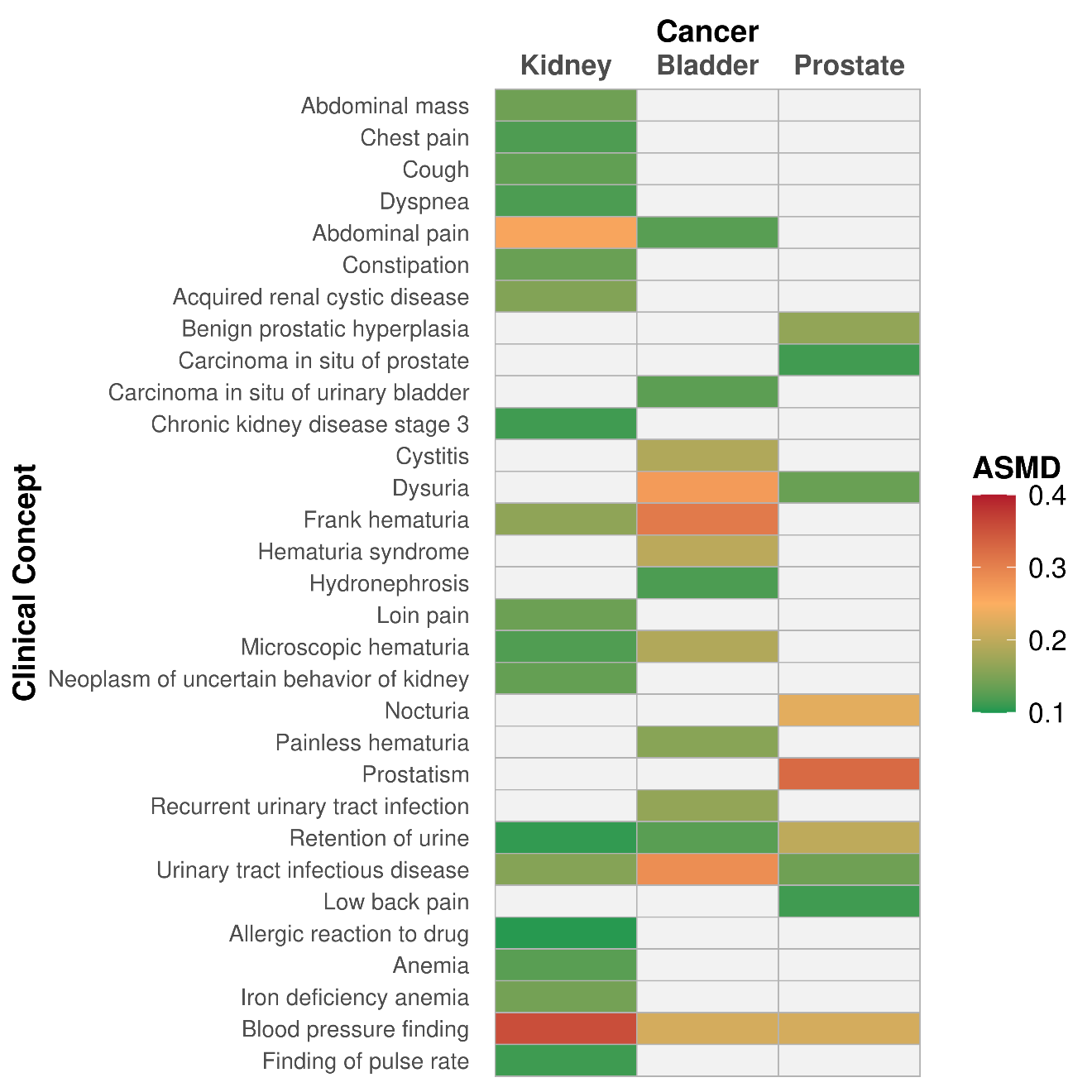

### **Figure S3: Site-specific clinical concepts with standardized mean differences (SMD) > 0.1 for female specific and breast cancers. Colours represent the magnitude of the SMD, with higher values indicating greater differences between cases and matched individuals.**

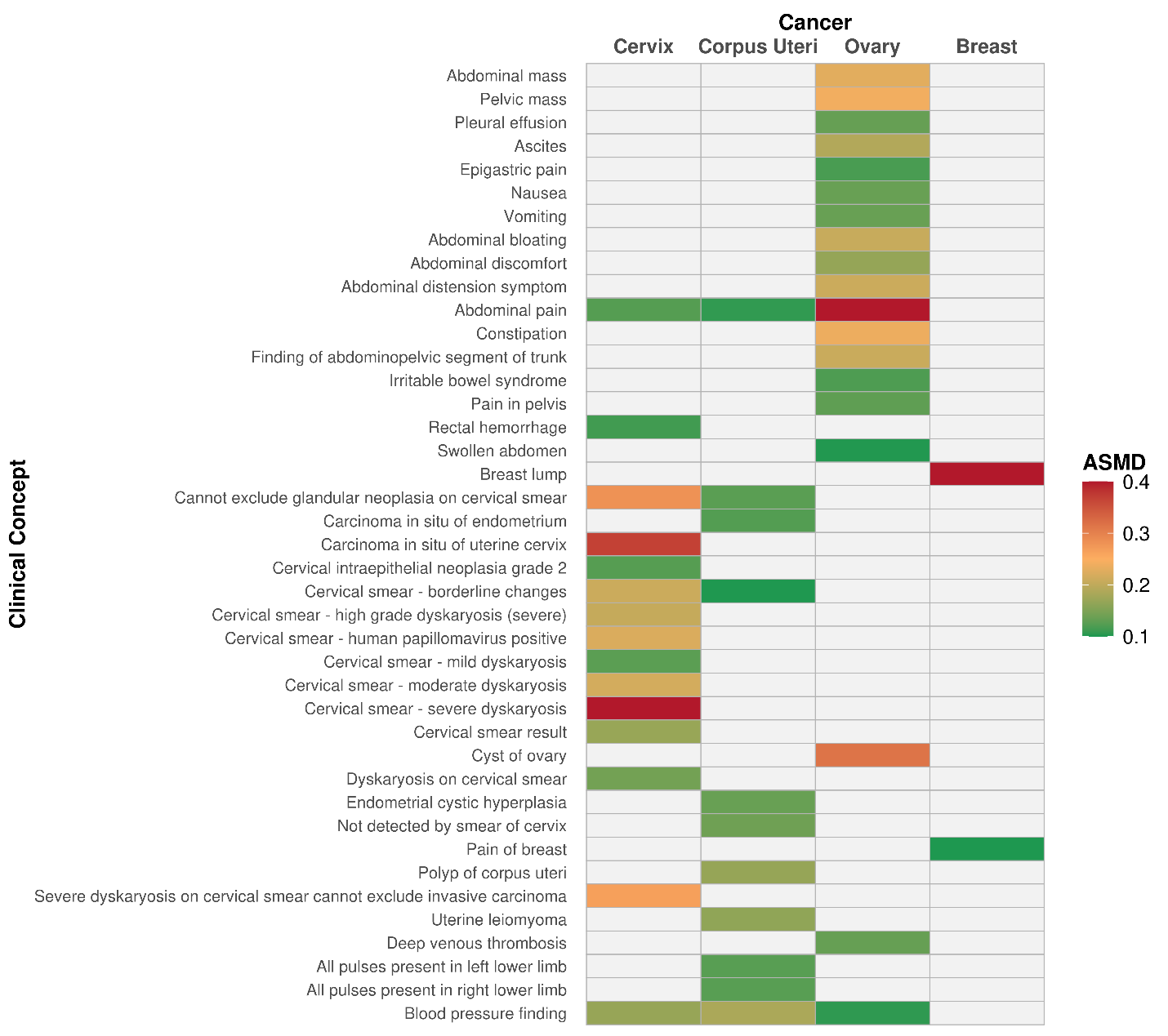

### **Figure S4: Site-specific clinical concepts with absolute standardized mean differences (SMD) > 0.1 for brain and thyroid cancers. Colours represent the magnitude of the SMD, with higher values indicating greater differences between cases and matched individuals.**

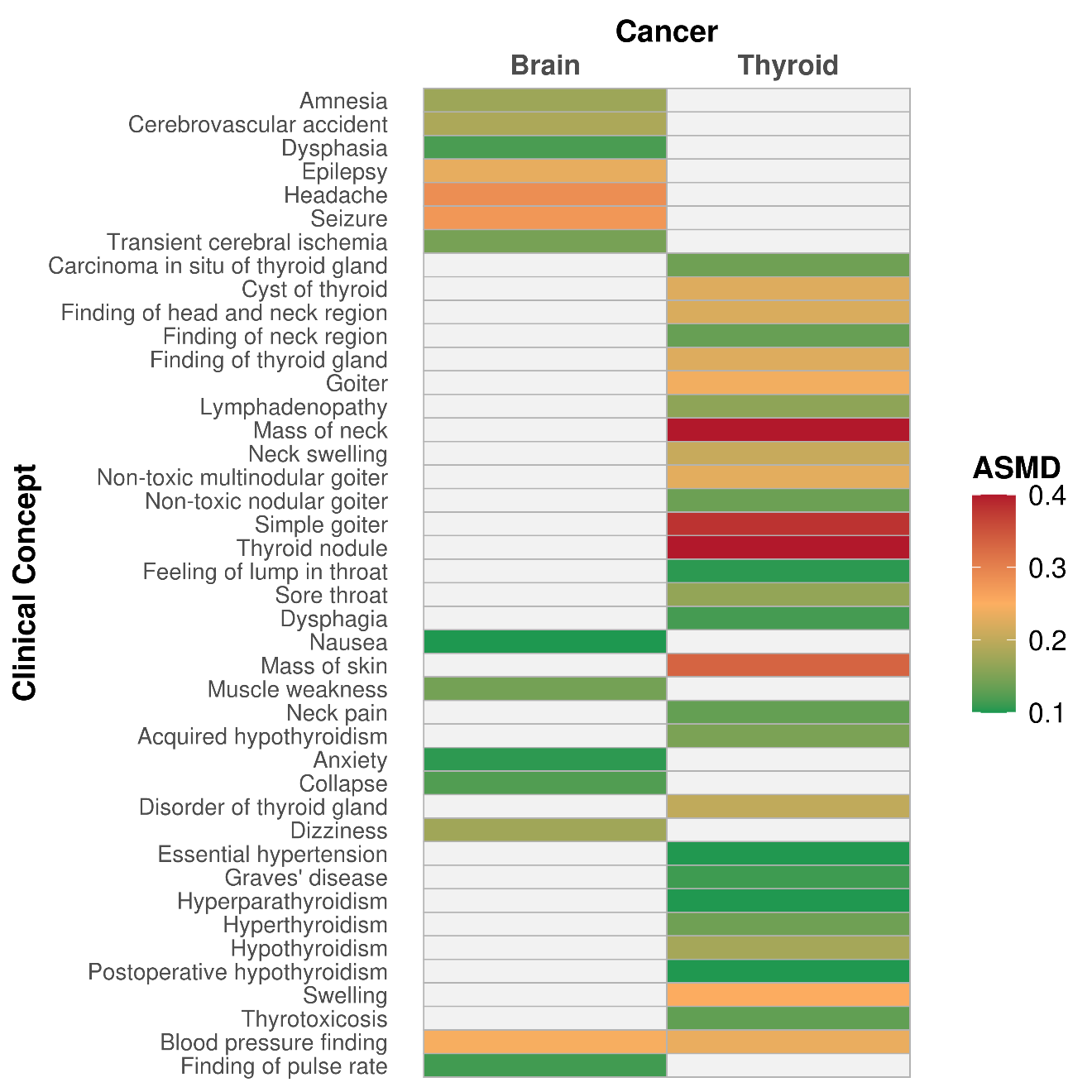

### **Table S3: Clinical concepts with absolute standardised mean differences (SMDs) > 0.1 for window 365-91 days before index showing percentages of each concept in cancer patients and matched individuals with the designation into flag group (RED SMD > 0.4, ORANGE SMD > 0.2 and <= 0.4, YELLOW SMD >0.1 and <= 0.2) and classification each presentation domain adapted from Zakkak *et al* (2024)**

| **Cancer** | **Code Name (Code ID)** | **Cancer %** | **Matched %** | **SMD** | **Assigned Domain** | **Assigned Flag** |
| --- | --- | --- | --- | --- | --- | --- |
| Anus | Abdominal pain (200219) | 3.03 | 1.44 | 0.1077 | Lower abdominal | YELLOW |
|  | Anal fissure (76725) | 1.74 | 0.15 | 0.1649 | Lower abdominal | YELLOW |
|  | Anal pain (442764) | 1.82 | 0 | 0.1925 | Lower abdominal | YELLOW |
|  | Anal skin tag (4112182) | 0.76 | 0 | 0.1238 | Lump/mass/lymph node | YELLOW |
|  | Atrophic vaginitis (201078) | 0.53 | 0 | 0.1032 | Female specific/breast | YELLOW |
|  | Constipation (75860) | 4.02 | 1.74 | 0.1366 | Lower abdominal | YELLOW |
|  | Diarrhea symptom (4145808) | 2.12 | 0.61 | 0.1304 | Lower abdominal | YELLOW |
|  | Diverticular disease (4266809) | 1.67 | 0.3 | 0.1391 | Lower abdominal | YELLOW |
|  | External hemorrhoids without complication (199839) | 2.81 | 0.08 | 0.2303 | Lower abdominal | ORANGE |
|  | Hemorrhoids (195562) | 8.26 | 0.45 | 0.3899 | Lower abdominal | ORANGE |
|  | Hiatal hernia (4224004) | 0.76 | 0.08 | 0.1053 | Upper abdominal | YELLOW |
|  | Internal hemorrhoids grade I (36715916) | 1.36 | 0 | 0.1661 | Lower abdominal | YELLOW |
|  | Painful rectal bleeding (4189790) | 0.99 | 0 | 0.1414 | Lower abdominal | YELLOW |
|  | Perianal abscess (199861) | 0.68 | 0 | 0.117 | Lower abdominal | YELLOW |
|  | Polyp of colon (4285898) | 0.76 | 0 | 0.1238 | Lower abdominal | YELLOW |
|  | Pruritus ani (200169) | 0.99 | 0.15 | 0.1118 | Lower abdominal | YELLOW |
|  | Rectal hemorrhage (4026112) | 6.37 | 0.3 | 0.343 | Lower abdominal | ORANGE |
|  | Rectal pain (76458) | 0.76 | 0 | 0.1238 | Lower abdominal | YELLOW |
|  | Rectal polyp (4216644) | 0.76 | 0.08 | 0.1053 | Lower abdominal | YELLOW |
|  | Thrombosed external hemorrhoids (201045) | 0.91 | 0.08 | 0.1185 | Lower abdominal | YELLOW |
| Bladder | Blood pressure finding (4214962) | 63.61 | 56.48 | 0.146 | Other clinical findings | YELLOW |
|  | Cystitis (195588) | 2.43 | 0.68 | 0.1418 | Genitourinary | YELLOW |
|  | Dysuria (197684) | 3.48 | 0.55 | 0.2097 | Genitourinary | ORANGE |
|  | Frank hematuria (75004) | 1.55 | 0.05 | 0.169 | Genitourinary | YELLOW |
|  | Hematuria syndrome (79864) | 0.9 | 0.05 | 0.1239 | Genitourinary | YELLOW |
|  | Microscopic hematuria (78472) | 0.89 | 0.03 | 0.1274 | Genitourinary | YELLOW |
|  | Recurrent urinary tract infection (4056621) | 1.2 | 0.18 | 0.1235 | Genitourinary | YELLOW |
|  | Urinary tract infectious disease (81902) | 4.89 | 1.24 | 0.213 | Genitourinary | ORANGE |
| Brain | Blood pressure finding (4214962) | 52.91 | 46.24 | 0.1337 | Other clinical findings | YELLOW |
|  | Cerebrovascular accident (381316) | 1 | 0.16 | 0.1108 | Central nervous system | YELLOW |
|  | Epilepsy (380378) | 1.51 | 0.1 | 0.1583 | Central nervous system | YELLOW |
|  | Headache (378253) | 2.97 | 1.06 | 0.1362 | Central nervous system | YELLOW |
|  | Seizure (377091) | 1.79 | 0.05 | 0.183 | Central nervous system | YELLOW |
| Breast | Breast lump (80767) | 2.44 | 0.49 | 0.1628 | Female specific/breast | YELLOW |
| Cervix | Cannot exclude glandular neoplasia on cervical smear (4056545) | 1.42 | 0 | 0.1697 | Female specific/breast | YELLOW |
|  | Carcinoma in situ of uterine cervix (194611) | 2.69 | 0 | 0.2351 | Female specific/breast | ORANGE |
|  | Cervical smear - borderline changes (4056546) | 2.69 | 0.39 | 0.1876 | Female specific/breast | YELLOW |
|  | Cervical smear - high grade dyskaryosis (severe) (44810424) | 0.61 | 0 | 0.1108 | Female specific/breast | YELLOW |
|  | Cervical smear - human papillomavirus positive (44810559) | 1.57 | 0.04 | 0.1718 | Female specific/breast | YELLOW |
|  | Cervical smear - mild dyskaryosis (4155377) | 1.11 | 0.17 | 0.1181 | Female specific/breast | YELLOW |
|  | Cervical smear - moderate dyskaryosis (4152694) | 1.4 | 0.02 | 0.1649 | Female specific/breast | YELLOW |
|  | Cervical smear - severe dyskaryosis (4151556) | 6.58 | 0.02 | 0.3736 | Female specific/breast | ORANGE |
|  | Severe dyskaryosis on cervical smear cannot exclude invasive carcinoma (4059358) | 0.85 | 0 | 0.1309 | Female specific/breast | YELLOW |
| Colon | Abdominal pain (200219) | 6.37 | 1.6 | 0.2457 | Lower abdominal | ORANGE |
|  | Anemia (439777) | 1.73 | 0.36 | 0.135 | Non-specific | YELLOW |
|  | Blood pressure finding (4214962) | 60.01 | 54.5 | 0.1116 | Other clinical findings | YELLOW |
|  | Constipation (75860) | 3.56 | 1.48 | 0.133 | Lower abdominal | YELLOW |
|  | Diarrhea symptom (4145808) | 2.28 | 0.98 | 0.1028 | Lower abdominal | YELLOW |
|  | Iron deficiency anemia (436659) | 3.65 | 0.46 | 0.2263 | Non-specific | ORANGE |
|  | Microcytic hypochromic anemia (4262948) | 1.43 | 0.1 | 0.15310000000000001 | Non-specific | YELLOW |
|  | Polyp of colon (4285898) | 0.88 | 0.11 | 0.1099 | Lower abdominal | YELLOW |
|  | Rectal hemorrhage (4026112) | 3.51 | 0.4 | 0.2261 | Lower abdominal | ORANGE |
| Corpus uteri | All pulses present in left lower limb (37312545) | 3.53 | 1.73 | 0.1127 | Other clinical findings | YELLOW |
|  | All pulses present in right lower limb (37312544) | 3.5 | 1.72 | 0.1118 | Other clinical findings | YELLOW |
|  | Blood pressure finding (4214962) | 58.62 | 51.52 | 0.1431 | Other clinical findings | YELLOW |
| Esophagus | Barrett's esophagus (443344) | 1.04 | 0.13 | 0.1195 | Upper abdominal | YELLOW |
|  | Blood pressure finding (4214962) | 59.91 | 54.41 | 0.1113 | Other clinical findings | YELLOW |
|  | Dysphagia (31317) | 4.23 | 0.31 | 0.2655 | Upper abdominal | ORANGE |
|  | Epigastric pain (197381) | 1.48 | 0.33 | 0.1217 | Upper abdominal | YELLOW |
|  | Gastroesophageal reflux disease (318800) | 0.86 | 0.12 | 0.1061 | Upper abdominal | YELLOW |
| Hypopharynx | Acute exacerbation of chronic obstructive pulmonary disease (257004) | 2.03 | 0.14 | 0.1832 | Respiratory | YELLOW |
|  | Acute pharyngitis (25297) | 1.3 | 0 | 0.1623 | Respiratory | YELLOW |
|  | Alcohol dependence (435243) | 1.01 | 0 | 0.1428 | Non-specific | YELLOW |
|  | Cellulitis (435613) | 1.45 | 0.43 | 0.1059 | Skin Lesion | YELLOW |
|  | Chronic obstructive pulmonary disease (255573) | 1.45 | 0.14 | 0.1479 | Respiratory | YELLOW |
|  | Cough (254761) | 8.55 | 5.94 | 0.1008 | Respiratory | YELLOW |
|  | Cramp in lower limb (42710045) | 0.58 | 0 | 0.108 | Musculoskeletal | YELLOW |
|  | Dysphagia (31317) | 3.48 | 0.58 | 0.2067 | Upper abdominal | ORANGE |
|  | Feeling of lump in throat (4145818) | 0.87 | 0 | 0.1325 | Respiratory | YELLOW |
|  | Finding of pulse rate (4103199) | 11.59 | 8.41 | 0.1061 | Other clinical findings | YELLOW |
|  | Hand pain (4204199) | 0.72 | 0 | 0.1204 | Musculoskeletal | YELLOW |
|  | Hemoptysis (261687) | 1.45 | 0 | 0.1715 | Respiratory | YELLOW |
|  | Intermittent claudication (442774) | 0.58 | 0 | 0.108 | Musculoskeletal | YELLOW |
|  | Lower respiratory tract infection (4175297) | 4.78 | 2.32 | 0.1332 | Respiratory | YELLOW |
|  | Mass of neck (4100606) | 0.87 | 0 | 0.1325 | Lump/mass/lymph node | YELLOW |
|  | Pain in throat (259153) | 2.61 | 0.29 | 0.195 | Respiratory | YELLOW |
|  | Pain of ear (4103476) | 2.75 | 0.72 | 0.1559 | Respiratory | YELLOW |
|  | Productive cough -clear sputum (4060224) | 1.01 | 0.14 | 0.1153 | Respiratory | YELLOW |
|  | Respiratory tract infection (4170143) | 0.87 | 0.14 | 0.1031 | Respiratory | YELLOW |
|  | Sciatica (372409) | 1.16 | 0.29 | 0.1027 | Musculoskeletal | YELLOW |
|  | Sore throat (4147326) | 8.99 | 0.87 | 0.3818 | Respiratory | ORANGE |
|  | Swelling (4275722) | 1.74 | 0.29 | 0.145 | Non-specific | YELLOW |
|  | Transient cerebral ischemia (373503) | 0.58 | 0 | 0.108 | Central nervous system | YELLOW |
| Kidney | Abdominal pain (200219) | 4.55 | 1.69 | 0.1651 | Lower abdominal | YELLOW |
|  | Acquired renal cystic disease (194686) | 0.67 | 0.03 | 0.1085 | Genitourinary | YELLOW |
|  | Blood pressure finding (4214962) | 63.79 | 50.13 | 0.2785 | Other clinical findings | ORANGE |
|  | Cough (254761) | 7.86 | 5.13 | 0.1109 | Respiratory | YELLOW |
|  | Frank hematuria (75004) | 0.72 | 0.05 | 0.1083 | Genitourinary | YELLOW |
|  | Urinary tract infectious disease (81902) | 2.58 | 1.1 | 0.1103 | Genitourinary | YELLOW |
| Larynx | Acute laryngitis (30133) | 0.86 | 0.03 | 0.1249 | Respiratory | YELLOW |
|  | Acute lower respiratory tract infection (4307774) | 2.1 | 0.83 | 0.1059 | Respiratory | YELLOW |
|  | Chronic laryngitis (24970) | 0.52 | 0 | 0.1022 | Respiratory | YELLOW |
|  | Chronic obstructive pulmonary disease (255573) | 2 | 0.59 | 0.125 | Respiratory | YELLOW |
|  | Cough (254761) | 9 | 5.38 | 0.1405 | Respiratory | YELLOW |
|  | Dysphagia (31317) | 1.62 | 0.14 | 0.159 | Upper abdominal | YELLOW |
|  | Dysplasia of vocal cord (4147413) | 0.52 | 0 | 0.1022 | Respiratory | YELLOW |
|  | Dyspnea (312437) | 3.24 | 1.48 | 0.1161 | Respiratory | YELLOW |
|  | Pain in throat (259153) | 1.55 | 0.14 | 0.1545 | Respiratory | YELLOW |
|  | Productive cough (4102774) | 1.35 | 0.41 | 0.1008 | Respiratory | YELLOW |
|  | Sore throat (4147326) | 6.62 | 0.52 | 0.3333 | Respiratory | ORANGE |
| Lip/Oral cavity | Candidiasis of mouth (29735) | 0.77 | 0.03 | 0.1174 | Ulceration | YELLOW |
|  | Disorder of tongue (432941) | 1.82 | 0.14 | 0.1712 | Ulceration | YELLOW |
|  | Lichen planus (132703) | 0.94 | 0.06 | 0.125 | Skin Lesion | YELLOW |
|  | Lymphadenopathy (315085) | 0.77 | 0.06 | 0.1106 | Lump/mass/lymph node | YELLOW |
|  | Mass of neck (4100606) | 0.94 | 0.03 | 0.1313 | Lump/mass/lymph node | YELLOW |
|  | Mouth symptoms (4060063) | 0.66 | 0.06 | 0.1003 | Ulceration | YELLOW |
|  | Pain in throat (259153) | 1.07 | 0.22 | 0.1063 | Respiratory | YELLOW |
|  | Pain of ear (4103476) | 2.04 | 0.52 | 0.1355 | Respiratory | YELLOW |
|  | Painful mouth (4010025) | 0.69 | 0.06 | 0.1032 | Ulceration | YELLOW |
|  | Sore throat (4147326) | 3 | 0.88 | 0.1542 | Respiratory | YELLOW |
|  | Ulcer of mouth (4094986) | 2.17 | 0.11 | 0.195 | Ulceration | YELLOW |
|  | Ulcer on tongue (4278836) | 0.55 | 0 | 0.1052 | Ulceration | YELLOW |
| Liver | 10g monofilament sensation L foot abnormal (4214578) | 1.58 | 0.39 | 0.1207 | Other clinical findings | YELLOW |
|  | 10g monofilament sensation L foot normal (4215145) | 7.67 | 2.78 | 0.2211 | Other clinical findings | ORANGE |
|  | 10g monofilament sensation R foot normal (4214579) | 7.65 | 2.56 | 0.2328 | Other clinical findings | ORANGE |
|  | Abdominal pain (200219) | 4.1 | 1.2 | 0.1813 | Lower abdominal | YELLOW |
|  | Alcoholic cirrhosis (196463) | 0.87 | 0 | 0.1325 | Upper abdominal | YELLOW |
|  | All pulses present in left foot (37312546) | 4.91 | 1.44 | 0.1989 | Other clinical findings | YELLOW |
|  | All pulses present in left lower limb (37312545) | 6.41 | 2.74 | 0.1763 | Other clinical findings | YELLOW |
|  | All pulses present in right foot (37312547) | 4.91 | 1.5 | 0.1945 | Other clinical findings | YELLOW |
|  | All pulses present in right lower limb (37312544) | 6.39 | 2.7 | 0.1779 | Other clinical findings | YELLOW |
|  | Anemia (439777) | 1.07 | 0.14 | 0.1201 | Non-specific | YELLOW |
|  | Ascites (200528) | 1.36 | 0 | 0.1661 | Upper abdominal | YELLOW |
|  | Blood pressure finding (4214962) | 66.44 | 52.83 | 0.2801 | Other clinical findings | ORANGE |
|  | Chest pain (77670) | 3.04 | 1.28 | 0.1213 | Respiratory | YELLOW |
|  | Cirrhosis of liver (4064161) | 3.2 | 0.02 | 0.2547 | Upper abdominal | ORANGE |
|  | Cough (254761) | 7.83 | 5.35 | 0.1001 | Respiratory | YELLOW |
|  | Dyspnea (312437) | 3.41 | 1.68 | 0.11 | Respiratory | YELLOW |
|  | Esophageal varices (24966) | 1.54 | 0.02 | 0.1734 | Upper abdominal | YELLOW |
|  | Finding of foot region (4022924) | 3.29 | 1.46 | 0.1204 | Other clinical findings | YELLOW |
|  | Finding of pulse rate (4103199) | 14.09 | 9.88 | 0.1299 | Other clinical findings | YELLOW |
|  | Gastritis (201340) | 1.07 | 0.18 | 0.1131 | Upper abdominal | YELLOW |
|  | Iron deficiency anemia (436659) | 1.44 | 0.39 | 0.1104 | Non-specific | YELLOW |
|  | Lower respiratory tract infection (4175297) | 3.77 | 2.07 | 0.1011 | Respiratory | YELLOW |
|  | Nonproliferative retinopathy of left eye due to diabetes mellitus (37311254) | 1.58 | 0.41 | 0.1181 | Non-specific | YELLOW |
|  | Normal pulse in left dorsalis pedis artery (37311332) | 1.46 | 0.43 | 0.1066 | Other clinical findings | YELLOW |
|  | Normal pulse in right dorsalis pedis artery (37311331) | 1.4 | 0.41 | 0.1047 | Other clinical findings | YELLOW |
|  | Normal tactile sensation (37311136) | 2.23 | 0.89 | 0.1083 | Other clinical findings | YELLOW |
|  | Portal hypertension (192680) | 0.95 | 0.02 | 0.1342 | Upper abdominal | YELLOW |
| Lung | Acute exacerbation of chronic obstructive pulmonary disease (257004) | 2.56 | 0.52 | 0.1662 | Respiratory | YELLOW |
|  | Acute lower respiratory tract infection (4307774) | 3.86 | 1.48 | 0.148 | Respiratory | YELLOW |
|  | Blood pressure finding (4214962) | 61.56 | 55.85 | 0.1162 | Other clinical findings | YELLOW |
|  | Chest pain (77670) | 3.95 | 1.75 | 0.1325 | Respiratory | YELLOW |
|  | Chronic obstructive pulmonary disease (255573) | 3.27 | 0.66 | 0.1889 | Respiratory | YELLOW |
|  | Cough (254761) | 13.26 | 6.05 | 0.246 | Respiratory | ORANGE |
|  | Dyspnea (312437) | 6.14 | 2.28 | 0.1931 | Respiratory | YELLOW |
|  | Hemoptysis (261687) | 1.5 | 0.16 | 0.1481 | Respiratory | YELLOW |
|  | Lower respiratory tract infection (4175297) | 5.73 | 2.22 | 0.1804 | Respiratory | YELLOW |
|  | Productive cough (4102774) | 2.2 | 0.85 | 0.1103 | Respiratory | YELLOW |
| Nasopharynx | Acute sinusitis (260123) | 3.38 | 0.48 | 0.212 | Respiratory | ORANGE |
|  | Acute tonsillitis (24660) | 0.97 | 0 | 0.14 | Respiratory | YELLOW |
|  | Alcohol dependence (435243) | 0.72 | 0 | 0.1204 | Non-specific | YELLOW |
|  | Allergic reaction to drug (4166257) | 3.38 | 0.72 | 0.1885 | Non-specific | YELLOW |
|  | Allergic rhinitis (257007) | 0.72 | 0 | 0.1204 | Respiratory | YELLOW |
|  | Anxiety (441542) | 1.21 | 0.24 | 0.1145 | Non-specific | YELLOW |
|  | Bleeding from nose (4096682) | 3.86 | 0.72 | 0.2111 | Respiratory | ORANGE |
|  | Cervical lymphadenopathy (4133459) | 1.45 | 0 | 0.1715 | Lump/mass/lymph node | YELLOW |
|  | Chronic catarrhal rhinitis (4145495) | 0.72 | 0 | 0.1204 | Respiratory | YELLOW |
|  | Chronic rhinitis (259848) | 1.45 | 0.24 | 0.1325 | Respiratory | YELLOW |
|  | Chronic sinusitis (257012) | 1.21 | 0 | 0.1565 | Respiratory | YELLOW |
|  | Community acquired pneumonia (4293463) | 0.72 | 0 | 0.1204 | Respiratory | YELLOW |
|  | Cough (254761) | 7.97 | 2.9 | 0.225 | Respiratory | ORANGE |
|  | Diplopia (373474) | 0.72 | 0 | 0.1204 | Central nervous system | YELLOW |
|  | Discomfort in mouth (4150136) | 0.72 | 0 | 0.1204 | Ulceration | YELLOW |
|  | Dysfunction of eustachian tube (381877) | 2.17 | 0.48 | 0.1482 | Respiratory | YELLOW |
|  | Dysphagia (31317) | 1.45 | 0.24 | 0.1325 | Upper abdominal | YELLOW |
|  | Edema (433595) | 0.72 | 0 | 0.1204 | Non-specific | YELLOW |
|  | Epidermoid cyst of skin (137351) | 0.72 | 0 | 0.1204 | Skin Lesion | YELLOW |
|  | Eruption (140214) | 2.9 | 0.24 | 0.2152 | Skin Lesion | ORANGE |
|  | Finding of head and neck region (255919) | 0.72 | 0 | 0.1204 | Lump/mass/lymph node | YELLOW |
|  | Gastric reflux (4023183) | 0.97 | 0 | 0.14 | Upper abdominal | YELLOW |
|  | Gastritis (201340) | 0.97 | 0 | 0.14 | Upper abdominal | YELLOW |
|  | Headache (378253) | 4.59 | 0.97 | 0.2215 | Central nervous system | ORANGE |
|  | Hearing loss (377889) | 3.86 | 0.97 | 0.1891 | Respiratory | YELLOW |
|  | Hyperlipidemia (432867) | 0.72 | 0 | 0.1204 | Non-specific | YELLOW |
|  | Increased blood pressure (4086587) | 1.21 | 0.24 | 0.1145 | Non-specific | YELLOW |
|  | Infective otitis externa (381859) | 2.17 | 0 | 0.2106 | Respiratory | ORANGE |
|  | Inflammatory dermatosis (45766714) | 0.72 | 0 | 0.1204 | Skin Lesion | YELLOW |
|  | Insomnia (436962) | 2.17 | 0.72 | 0.1217 | Non-specific | YELLOW |
|  | Iron deficiency anemia (436659) | 0.72 | 0 | 0.1204 | Non-specific | YELLOW |
|  | Lower respiratory tract infection (4175297) | 3.14 | 1.45 | 0.113 | Respiratory | YELLOW |
|  | Lymphadenopathy (315085) | 2.66 | 0 | 0.2338 | Lump/mass/lymph node | ORANGE |
|  | Mass of neck (4100606) | 4.11 | 0 | 0.2928 | Lump/mass/lymph node | ORANGE |
|  | Mass of skin (4183953) | 2.17 | 0 | 0.2106 | Skin Lesion | ORANGE |
|  | Mouth symptoms (4060063) | 0.72 | 0 | 0.1204 | Ulceration | YELLOW |
|  | Nasal congestion (4195085) | 1.21 | 0 | 0.1565 | Respiratory | YELLOW |
|  | Nasal symptom (4091808) | 1.21 | 0.24 | 0.1145 | Respiratory | YELLOW |
|  | Non-suppurative otitis media with eustachian tube disorder (4111114) | 0.72 | 0 | 0.1204 | Respiratory | YELLOW |
|  | Normal pulse in left dorsalis pedis artery (37311332) | 0.97 | 0 | 0.14 | Other clinical findings | YELLOW |
|  | Normal pulse in right dorsalis pedis artery (37311331) | 0.97 | 0 | 0.14 | Other clinical findings | YELLOW |
|  | Normal tactile sensation (37311136) | 1.21 | 0.24 | 0.1145 | Other clinical findings | YELLOW |
|  | Not detected by smear of cervix (4155376) | 3.38 | 1.69 | 0.1077 | Female specific/breast | YELLOW |
|  | Pain in throat (259153) | 1.21 | 0 | 0.1565 | Respiratory | YELLOW |
|  | Pain of ear (4103476) | 3.38 | 0.48 | 0.212 | Respiratory | ORANGE |
|  | Posterior rhinorrhea (257683) | 0.72 | 0 | 0.1204 | Respiratory | YELLOW |
|  | Purulent otitis media (376712) | 0.72 | 0 | 0.1204 | Respiratory | YELLOW |
|  | Sinus headache (4172302) | 1.69 | 0 | 0.1854 | Respiratory | YELLOW |
|  | Sore throat (4147326) | 5.07 | 0.97 | 0.2413 | Respiratory | ORANGE |
|  | Swelling (4275722) | 1.93 | 0 | 0.1984 | Non-specific | YELLOW |
|  | Wax in ear canal (4155902) | 2.9 | 1.21 | 0.1193 | Respiratory | YELLOW |
| Oropharynx | Acute pharyngitis (25297) | 0.88 | 0.08 | 0.1159 | Respiratory | YELLOW |
|  | Acute tonsillitis (24660) | 1.38 | 0 | 0.1673 | Respiratory | YELLOW |
|  | Cervical lymphadenopathy (4133459) | 0.65 | 0 | 0.1144 | Lump/mass/lymph node | YELLOW |
|  | Dysphagia (31317) | 1.42 | 0.19 | 0.138 | Upper abdominal | YELLOW |
|  | Lymphadenopathy (315085) | 2.1 | 0.04 | 0.2012 | Lump/mass/lymph node | ORANGE |
|  | Mass of neck (4100606) | 3.25 | 0 | 0.2592 | Lump/mass/lymph node | ORANGE |
|  | Mass of skin (4183953) | 1.15 | 0.23 | 0.1113 | Skin Lesion | YELLOW |
|  | Pain in throat (259153) | 1.65 | 0.04 | 0.1766 | Respiratory | YELLOW |
|  | Pain of ear (4103476) | 3.18 | 0.61 | 0.1893 | Respiratory | YELLOW |
|  | Sore throat (4147326) | 6.2 | 0.69 | 0.3056 | Respiratory | ORANGE |
|  | Swelling (4275722) | 1.84 | 0.23 | 0.1596 | Non-specific | YELLOW |
| Ovary | Abdominal bloating (4023572) | 1.1 | 0.21 | 0.1105 | Lower abdominal | YELLOW |
|  | Abdominal discomfort (4183041) | 0.92 | 0.14 | 0.1076 | Lower abdominal | YELLOW |
|  | Abdominal distension symptom (4012079) | 0.72 | 0.06 | 0.106 | Lower abdominal | YELLOW |
|  | Abdominal mass (192438) | 0.68 | 0.01 | 0.1145 | Lump/mass/lymph node | YELLOW |
|  | Abdominal pain (200219) | 8.64 | 1.99 | 0.2997 | Lower abdominal | ORANGE |
|  | Constipation (75860) | 3.43 | 1.26 | 0.1438 | Lower abdominal | YELLOW |
|  | Cyst of ovary (197610) | 1.71 | 0.07 | 0.1753 | Female specific/breast | YELLOW |
|  | Pelvic mass (201131) | 0.57 | 0 | 0.1071 | Lump/mass/lymph node | YELLOW |
| Pancreas | 10g monofilament sensation L foot normal (4215145) | 4.79 | 2.51 | 0.1218 | Other clinical findings | YELLOW |
|  | 10g monofilament sensation R foot normal (4214579) | 4.9 | 2.43 | 0.1317 | Other clinical findings | YELLOW |
|  | Abdominal pain (200219) | 8.27 | 1.87 | 0.2949 | Lower abdominal | ORANGE |
|  | All pulses present in left lower limb (37312545) | 4.61 | 2.5 | 0.1141 | Other clinical findings | YELLOW |
|  | All pulses present in right lower limb (37312544) | 4.63 | 2.48 | 0.1163 | Other clinical findings | YELLOW |
|  | Blood pressure finding (4214962) | 63.49 | 55.08 | 0.1718 | Other clinical findings | YELLOW |
|  | Constipation (75860) | 4.06 | 1.63 | 0.1466 | Lower abdominal | YELLOW |
|  | Diabetes mellitus (201820) | 2.14 | 0.42 | 0.1535 | Upper abdominal | YELLOW |
|  | Epigastric pain (197381) | 3.03 | 0.3 | 0.2146 | Upper abdominal | ORANGE |
|  | Jaundice (137977) | 1.62 | 0.04 | 0.1748 | Upper abdominal | YELLOW |
|  | Obstructive hyperbilirubinemia (4242900) | 1.11 | 0.02 | 0.1458 | Upper abdominal | YELLOW |
|  | Type 2 diabetes mellitus (201826) | 4.51 | 1.1 | 0.2076 | Upper abdominal | ORANGE |
| Prostate | Benign prostatic hyperplasia (198803) | 1.75 | 0.46 | 0.1236 | Genitourinary | YELLOW |
|  | Blood pressure finding (4214962) | 61.9 | 55.58 | 0.1286 | Other clinical findings | YELLOW |
|  | Nocturia (40304526) | 2.39 | 0.61 | 0.1468 | Genitourinary | YELLOW |
|  | Prostatism (4016155) | 4.02 | 0.7 | 0.22 | Genitourinary | ORANGE |
|  | Retention of urine (192450) | 1.74 | 0.29 | 0.145 | Genitourinary | YELLOW |
|  | Urinary tract infectious disease (81902) | 2.01 | 0.8 | 0.1029 | Genitourinary | YELLOW |
| Rectum | Constipation (75860) | 3.72 | 1.47 | 0.1419 | Lower abdominal | YELLOW |
|  | Diarrhea (196523) | 1.26 | 0.34 | 0.1034 | Lower abdominal | YELLOW |
|  | Diarrhea symptom (4145808) | 2.98 | 0.84 | 0.1568 | Lower abdominal | YELLOW |
|  | Hemorrhoids (195562) | 2.42 | 0.4 | 0.172 | Lower abdominal | YELLOW |
|  | Rectal hemorrhage (4026112) | 6.53 | 0.38 | 0.3416 | Lower abdominal | ORANGE |
|  | Rectal polyp (4216644) | 0.77 | 0.03 | 0.1174 | Lower abdominal | YELLOW |
| Salivary gland | Adenolymphoma (4307552) | 0.74 | 0 | 0.1221 | Lump/mass/lymph node | YELLOW |
|  | Basal cell carcinoma of skin (4112752) | 1.72 | 0.25 | 0.1493 | Skin Lesion | YELLOW |
|  | Bell's palsy (374923) | 2.58 | 0 | 0.2301 | Central nervous system | ORANGE |
|  | Blood pressure finding (4214962) | 53.44 | 48.28 | 0.1034 | Other clinical findings | YELLOW |
|  | Cervical lymphadenopathy (4133459) | 0.61 | 0 | 0.1108 | Lump/mass/lymph node | YELLOW |
|  | Diarrhea symptom (4145808) | 1.23 | 0.25 | 0.1145 | Lower abdominal | YELLOW |
|  | Eruption (140214) | 2.83 | 0.98 | 0.1356 | Skin Lesion | YELLOW |
|  | Eye symptom (4205680) | 0.98 | 0.12 | 0.1165 | Non-specific | YELLOW |
|  | Finding of lymph node (4200648) | 0.61 | 0 | 0.1108 | Lump/mass/lymph node | YELLOW |
|  | Itching (4302654) | 0.61 | 0 | 0.1108 | Non-specific | YELLOW |
|  | Jaw pain (4168686) | 0.86 | 0.12 | 0.1061 | Non-specific | YELLOW |
|  | Lymphadenopathy (315085) | 3.44 | 0 | 0.2669 | Lump/mass/lymph node | ORANGE |
|  | Mass of neck (4100606) | 4.42 | 0.12 | 0.2918 | Lump/mass/lymph node | ORANGE |
|  | Mass of parotid gland (4202556) | 2.09 | 0 | 0.2066 | Lump/mass/lymph node | ORANGE |
|  | Mass of skin (4183953) | 5.41 | 0 | 0.3382 | Skin Lesion | ORANGE |
|  | Pain in face (4317968) | 0.74 | 0 | 0.1221 | Non-specific | YELLOW |
|  | Pain of ear (4103476) | 3.07 | 0.74 | 0.1711 | Respiratory | YELLOW |
|  | Palpitations (315078) | 1.11 | 0.25 | 0.1048 | Non-specific | YELLOW |
|  | Parotid swelling (4152183) | 3.93 | 0.12 | 0.273 | Lump/mass/lymph node | ORANGE |
|  | Parotitis (4032796) | 0.86 | 0 | 0.1317 | Non-specific | YELLOW |
|  | Skin lesion (4316083) | 2.09 | 0.74 | 0.1145 | Skin Lesion | YELLOW |
|  | Sore throat (4147326) | 2.09 | 0.61 | 0.1285 | Respiratory | YELLOW |
|  | Squamous cell carcinoma of skin (4111921) | 0.86 | 0 | 0.1317 | Skin Lesion | YELLOW |
|  | Swelling (4275722) | 6.88 | 0.61 | 0.3348 | Non-specific | ORANGE |
| Stomach | Abdominal pain (200219) | 5.47 | 1.94 | 0.1877 | Lower abdominal | YELLOW |
|  | Anemia (439777) | 1.59 | 0.23 | 0.1436 | Non-specific | YELLOW |
|  | Blood pressure finding (4214962) | 64.54 | 58.36 | 0.1272 | Other clinical findings | YELLOW |
|  | Chest pain (77670) | 4.18 | 1.9 | 0.1331 | Respiratory | YELLOW |
|  | Constipation (75860) | 3.68 | 1.52 | 0.136 | Lower abdominal | YELLOW |
|  | Dysphagia (31317) | 1.24 | 0.34 | 0.1018 | Upper abdominal | YELLOW |
|  | Epigastric pain (197381) | 3.15 | 0.38 | 0.2115 | Upper abdominal | ORANGE |
|  | Gastric ulcer (4265600) | 1.43 | 0.09 | 0.1548 | Upper abdominal | YELLOW |
|  | Iron deficiency anemia (436659) | 3.06 | 0.4 | 0.2051 | Non-specific | ORANGE |
|  | Microcytic hypochromic anemia (4262948) | 1.25 | 0.09 | 0.1426 | Non-specific | YELLOW |
|  | Vomiting (441408) | 1.67 | 0.48 | 0.1156 | Upper abdominal | YELLOW |
| Thyroid | Blood pressure finding (4214962) | 49.06 | 39.63 | 0.1907 | Other clinical findings | YELLOW |
|  | Cyst of thyroid (138113) | 2.13 | 0 | 0.2086 | Lump/mass/lymph node | ORANGE |
|  | Disorder of thyroid gland (141253) | 1.13 | 0 | 0.1512 | Non-specific | YELLOW |
|  | Finding of head and neck region (255919) | 1.54 | 0 | 0.1769 | Lump/mass/lymph node | YELLOW |
|  | Finding of thyroid gland (4201376) | 1.95 | 0 | 0.1994 | Lump/mass/lymph node | YELLOW |
|  | Goiter (135772) | 2.09 | 0 | 0.2066 | Lump/mass/lymph node | ORANGE |
|  | Graves' disease (4232076) | 0.51 | 0 | 0.1013 | Non-specific | YELLOW |
|  | Hyperthyroidism (4142479) | 0.86 | 0 | 0.1317 | Non-specific | YELLOW |
|  | Hypothyroidism (140673) | 1.58 | 0.34 | 0.1274 | Non-specific | YELLOW |
|  | Lymphadenopathy (315085) | 0.93 | 0.1 | 0.1162 | Lump/mass/lymph node | YELLOW |
|  | Mass of neck (4100606) | 8.06 | 0.1 | 0.4108 | Lump/mass/lymph node | RED |
|  | Mass of skin (4183953) | 3.91 | 0.17 | 0.2669 | Skin Lesion | ORANGE |
|  | Neck pain (24134) | 2.13 | 0.69 | 0.1224 | Musculoskeletal | YELLOW |
|  | Neck swelling (4200520) | 1.3 | 0 | 0.1623 | Lump/mass/lymph node | YELLOW |
|  | Non-toxic multinodular goiter (136368) | 1.99 | 0 | 0.2015 | Lump/mass/lymph node | ORANGE |
|  | Non-toxic nodular goiter (136661) | 0.79 | 0 | 0.1262 | Lump/mass/lymph node | YELLOW |
|  | Simple goiter (141825) | 5.31 | 0 | 0.3349 | Lump/mass/lymph node | ORANGE |
|  | Sore throat (4147326) | 3.36 | 1.51 | 0.1202 | Respiratory | YELLOW |
|  | Swelling (4275722) | 2.98 | 0.31 | 0.2111 | Non-specific | ORANGE |
|  | Thyroid nodule (4030041) | 6.48 | 0 | 0.3723 | Lump/mass/lymph node | ORANGE |
|  | Thyrotoxicosis (138387) | 0.89 | 0.07 | 0.1189 | Non-specific | YELLOW |
